# Ensemble forecasting of influenza activity and assessing its year-round dynamical characteristics during and post-COVID-19 pandemic periods in a sub-tropical location

**DOI:** 10.64898/2025.12.17.25342517

**Authors:** Dong Wang, Yiu-Chung Lau, Songwei Shan, Dongxuan Chen, Zhanwei Du, Eric H. Y. Lau, Daihai He, Linwei Tian, Peng Wu, Benjamin J. Cowling, Sheikh Taslim Ali

## Abstract

Influenza forecasting in (sub-)tropical regions remains understudied due to year-round, irregular transmission patterns. Further, the variation in seasonality and transmission characteristic of influenza in post-COVID-19 pandemic could be attributed to various drivers to quantify for better understanding. To address this issue, this study introduced an ensemble forecasting approach that incorporates varied dataset lengths to forecast influenza activity in Hong Kong, integrating multi-stream surveillance data, including absolute humidity, temperature, ozone, and school closures/holidays. We applied temporal cross-validation to evaluate forecasting performance for short- and long term separately across different training-sets and model variants, ultimately constructing ensemble forecasts weighted by individual model performance. The optimal ensemble model could forecast the 2019/20 winter influenza season onwards and evaluate the impact of COVID-19 public health and social measures (PHSMs). We further extended the framework to forecast influenza in post-pandemic period since March 2023, accounting for the impact of cessation of PHSMs and COVID-19-induced cross-protection/competition in population susceptibility. Forecasts showed two peaks in 2019/20 season, which could account for 95.2% (95% prediction interval (PI): 89.1%, 98.3%) reduction in attack rate for COVID-19 PHSMs. The post-pandemic forecasts indicated changes in influenza transmission dynamics and seasonality, highlighting the need to consider factors such as population immunity and co-circulation with COVID-19 in future influenza forecasts. This study emphasizes the importance of incorporating diverse factors for better influenza forecasts in (sub-)tropical regions. The proposed framework offers a scalable tool for forecasting other respiratory virus transmissions, supporting healthcare agencies in managing future infection burdens and enhancing preparedness.

**Author summary:** Reliable and proactive forecasts of influenza activity and timing of epidemic outcomes enable public health officials to plan targeted responses. However, unlike temperate locations, the irregular seasonality of influenza in tropical/subtropical locations leads to highly variable forecasting patterns when models use varying lengths of historical data, reducing the robustness of forecasts. By leveraging multi-stream surveillance data in Hong Kong, we developed a mechanistic model-based ensemble forecasting framework that integrate potential combinations of data and models for short-, medium-, and long-term forecasts of influenza outcomes. Beyond methodological advancement, this framework has broader implications in assessing the impact of COVID-19-related interventions on influenza dynamics during pandemic and evaluating potential co-circulation risk of respiratory viruses including influenza and COVID-19 in post-pandemic era.

## INTRODUCTION

Influenza, a recurring global health concern, is responsible for an estimated 300,000 to 650,000 deaths annually, highlighting its significant public health burden ^1,2^. Modeling and forecasting influenza transmission dynamics offer valuable insights into epidemic behavior and hold the potential to improve preparedness for future outbreaks, with profound implications for public health and individual lives^3–6^. In temperate regions, such as the continental United States, influenza transmission follows a distinct seasonal pattern ^7,8^. Reliable forecasting in these regions has been achieved with the leverage of numerous methods, including statistical, mechanistic, and machine learning models ^3,9–16^. However, the scenarios differ markedly in tropical and subtropical regions, where epidemic patterns are more intricate and less regular, often featuring multiple waves ^7,8,17–19^. Influenza epidemics occur throughout the year, with peaks in the winter almost every year and secondary peaks in the spring, summer, or autumn in some years ^18,19^. The year-round less regular epidemic dynamics, coupled with the lack of real-time information on the epidemic outcomes, make predicting and forecasting influenza dynamics in (sub-)tropical regions a challenging task ^20–22^.

Several environmental, climatic and social factors, including absolute humidity (AH) ^23,24^, temperature^25–28^, ozone levels ^29–31^, and school closures & holidays ^32–35^ could drive the influenza transmission in community. In temperate locations, low absolute humidity in winter often coincides with an increase in influenza cases ^24^. However, in subtropical regions, the pattern is less predictable, exhibiting a “U-shape” relationship between transmissibility and absolute humidity ^26,36,37^. While lower temperature can increase virus survival and transmission, potentially leading to more deaths among vulnerable groups ^27^. Ambient ozone concertation could potentially reduce infectivity of influenza virus ^29–31^. The opening and closure of schools can affect influenza dynamics, although the impact on transmission isn’t always straightforward ^32–35^. Public Health and Social Measures (PHSMs), which have been widely used to curb COVID-19, could also affect influenza transmission^38–42^.

Mechanistic models offer an effective method for understanding disease dynamics ^5,14^. However, their forecasting performance can vary based on the size of the historical data used and the approach to incorporate these drivers of influenza in the models. To account for these uncertainties and potential deviations, ensemble models, which combine different models, have been proposed ^6,10,13,43^. This approach leverages the strengths of each model and can often result in more accurate and reliable forecasts ^43,44^. It’s worth noting that the choice of forecasting model and method depends on several key characteristics, including the nature of the data, the forecast horizon, and the accuracy requirements of the forecast ^10^. Ensemble forecasting avoids relying solely on the assumptions and hypotheses of a single model, leading to more reliable and robust forecasts of influenza activity, even with comparatively less regular seasonality ^43–45^.

Therefore, we performed ensemble forecasting of influenza activity in Hong Kong, a subtropical city, using multi-stream surveillance data, including syndromic, virological, and extrinsic drivers of influenza (e.g., absolute humidity, temperature, ozone levels, school closures, and holidays). To capture the year-round dynamics of influenza transmission in Hong Kong, we developed 16 models, each incorporating different combinations of these drivers within the time-space modeling framework (e.g., the Susceptible-Vaccination-Exposed-Infectious-Recovered-Susceptible (SVEIRS) model). We evaluated the forecasting performance of each model using varying lengths of historical data through temporal cross-validation and combined them into an ensemble forecast using a rank-based weighting method to increase the reliability of our forecast. We implemented our framework to evaluate the impact of COVID-19 pandemic and assessed the dynamical characteristics of influenza during and post-pandemic periods in Hong Kong.

## RESULTS

### Influenza activity in Hong Kong

In Hong Kong, the influenza activity (ILI proxy) since 2010 showed less regular, with a primary winter-spring peak and a secondary peak often occurring in summer (**Fig. 1a)**. During the 2019/20 season, influenza activity sharply declined after the first COVID-19 case was reported on 22^nd^ January, due to the implementation of PHSMs. This low activity persisted until the 2022/23 winter season as these measures remained in place to mitigate COVID-19. In the post-pandemic period, upon relaxation of these PHSMs on 1^st^ Mar 2023, the influenza activity reemerged in the 2023 spring season and exhibited noticeable changes in seasonal patterns, including variations in peak timing and magnitude. The successive summer season in 2023 was delayed with a peak in late September and a relatively low activity during the following 2023-24 winter season (**Fig. 1a)**.

**Figure 1.**
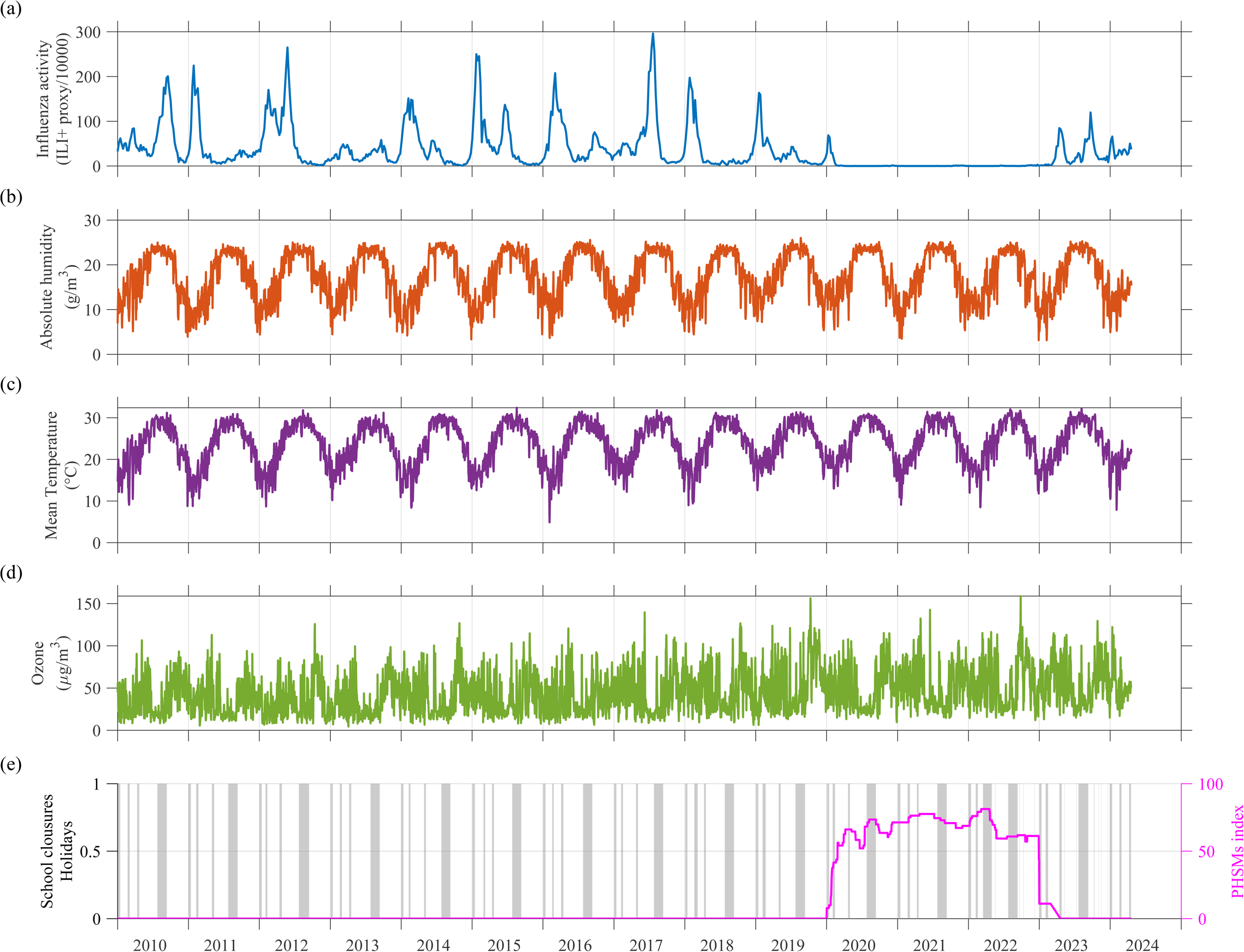
Observed influenza epidemics and surveillance data in Hong Kong from January 2010 to April 2024. (a) Weekly ILI+ proxy (per 10^5^ population) to represent the influenza activity in Hong Kong. Daily mean (b) absolute humidity, (c) temperature, and (d) ambient ozone concentration, and (e) the timing and duration of school closures and holidays during the study period in gray bars, and the PHSMs index time series from the Oxford Covid-19 Government Response Tracker (OxCGRT) in pink line.

### Model forecasting performance under different ensemble schemes

We used multi-stream data, including absolute humidity, temperature, ozone concentration, and school closures/holidays, to characterize seasonal transmission rates for developing SVEIRS-type models of influenza in Hong Kong (**Fig S1** and Supplementary Section 1). By considering all possible combinations of these drivers, we constructed 16 individual model variants and 5 ensemble models (MATERIALS AND METHODS). We used temporal cross-validation methods and weighted interval scores (WIS) to evaluate their forecast performance on windows of 7, 13, 26, and 52 weeks during the validation period of 1^st^ January 2017 – 31^st^ December 2019 (MATERIALS AND METHODS). The WIS of individual model variants and ensemble models were illustrated in **Fig. 2** and **Fig. 3**, respectively. The temporal cross-validation revealed that short-term (up to 7–13 weeks ahead), medium-term (up to 26 weeks ahead) and long-term (up to 52 weeks ahead) forecasting performance may vary depending on the historical training datasets and model variants. Generally, individual models trained on shorter (1–3 years), medium (2–4 years), and longer (4–7 years) lengths of historical data performed well for short-term, medium-term, and long-term forecasting, respectively (**Fig. 2**). We found that the ensemble model incorporating all potential drivers (in model variant E-IV) exhibited the best performance across various forecasting windows (**Fig. 3**).

**Figure 2.**
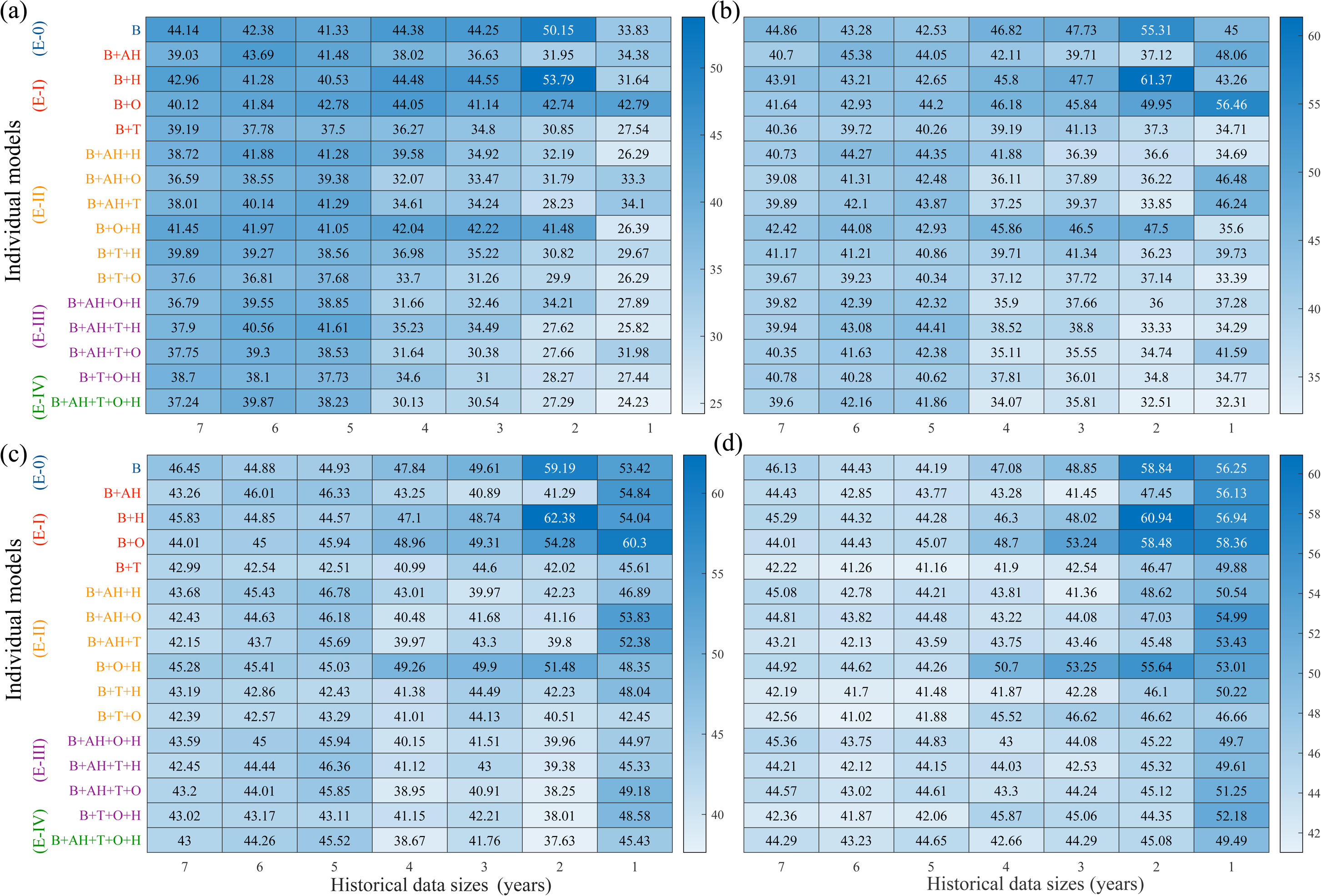
The Weighted Interval Score (WIS) of the candidate individual models, trained using different lengths of historical data (1 - 7 years) for different forecasting windows - (a) 7 weeks ahead, (b) 13 weeks ahead, (c) 26 weeks ahead, (d) 52 weeks ahead. Smaller WIS values indicate better models. The y-axis labels with different colors represent the respective individual models under the same ensemble model (E-0, E-I, E-II, E-III, and E-IV). E-0, E-I, E-II, E-III, and E-IV indicate the respective ensemble models aggregated by up to 0 to 4 factors in individual models, where 0 factor indicates the base-model (E-0 or B). Further, each of these models was trained by using 7 different lengths (1, 2,…, and 7 years) of historical data windows.

**Figure 3.**
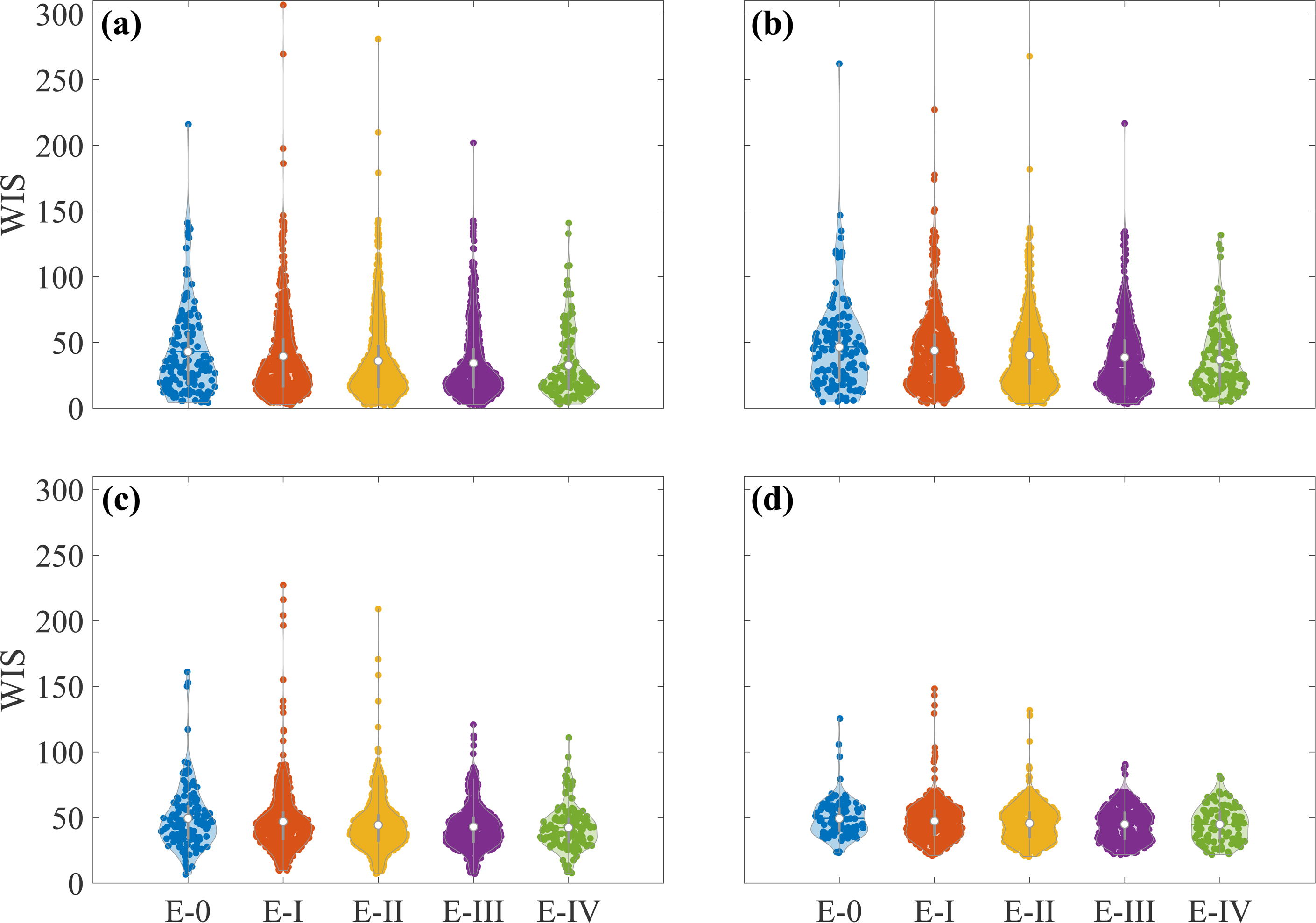
Forecasting performance (based on WIS) among 5 ensemble models, for different forecasting terms - (a) 7 weeks ahead, (b) 13 weeks ahead, (c) 26 weeks ahead, (d) 52 weeks ahead. The violin represents the distributions of the WIS values for each individual model candidate under each cross-validation (in dots), which were used to construct ensemble model. The medium of these WIS values indicates the forecasting performance for the respective ensemble model.

### Impact of COVID-19 pandemic on influenza activity since January 2020

For short-term forecasting, we examined up to 7-week ahead forecasts starting from 22^nd^ January 2020, which showed an increasing trend in influenza activity until late February and then started to decline (**Fig. 4a**). We projected that the peak could occur during the week of 23^rd^-29^th^ February, with a peak magnitude of 94.79 (95% PI: 31.59,165.99) per 10,000 consultations. During the 7 weeks (22^nd^ January to 11^th^ March), the predicted attack rates could be 5.49% (95% PI: 1.51%, 9.21%), accounting for an 81.28% (95% PI: 45.35%, 91.02%) reduction in influenza attack rates when compared to the observed attack rates under PHSMs. With up to a 13-week ahead forecast window, we found a similar trend of influenza activity with peak timing and peak magnitude (**Fig. 4b**), predicting an attack rate of 7.95% (95% PI: 3.28%, 13.51%), which is associated with a reduction of 87.76% (95% PI: 73.94%, 93.68%) in attack rates (**Table 1**). For medium-term forecasts, it exhibited influenza activity with a projected resurgence in late April that could be sustained until July 2020, mimicking the usual summer season with a second peak in the week of 5^th^ −11^th^ July 2020, accounting for a reduction of 93.49% (95% PI: 85.69%, 97.66%) in attack rates (**Fig. 4c**). Meanwhile, for long-term forecasting, up to a year’s forecast predicted two clear peaks during the pandemic period with a spring peak in early March and a summer peak in July, similar to the medium-term forecast peak timings. We observed a week of delay in the first peak onsets under medium and long-term compared to short-term forecasting. The respective peak magnitudes were comparable with tiny variations and accounted for up to a 95.15% (95% PI: 89.07%, 98.29%) reduction in attack rates for the entire year (**Fig. 4d and Table 1)**. To assess the sensitivity of forecasting start date, we evaluated two event-based forecasting start dates: January 6th (when airport and train station screenings began for travelers from Wuhan) and January 15th (the week preceding first confirmed COVID-19 case in Hong Kong). The results for these dates across various periods were shown in **Figs. S2-3**. Our analysis found that the choice of start date exerted minimal impact on influenza activity forecasts (Supplementary Section 2). In addition, we observed the forecasting patterns could vary with different lengths of historical datasets (1-7 years) for long-term forecasting starting from January 22, 2020 (**Fig. 5)**, therefore, underscoring the rationale for adopting ensemble forecasting. The forecasting results showed that longer length of historical data (5-7 years) could capture long-term seasonal patterns and reflect the forecasting trends better. Medium length of historical data (3-4 years) could compromise in detecting long-term seasonality and could capture short-term fluctuations while displaying comparable forecasting patterns. The shorter length of historical data (1-2 years) could focus more on recent changes and return better short-term forecasting trends.

**Figure 4.**
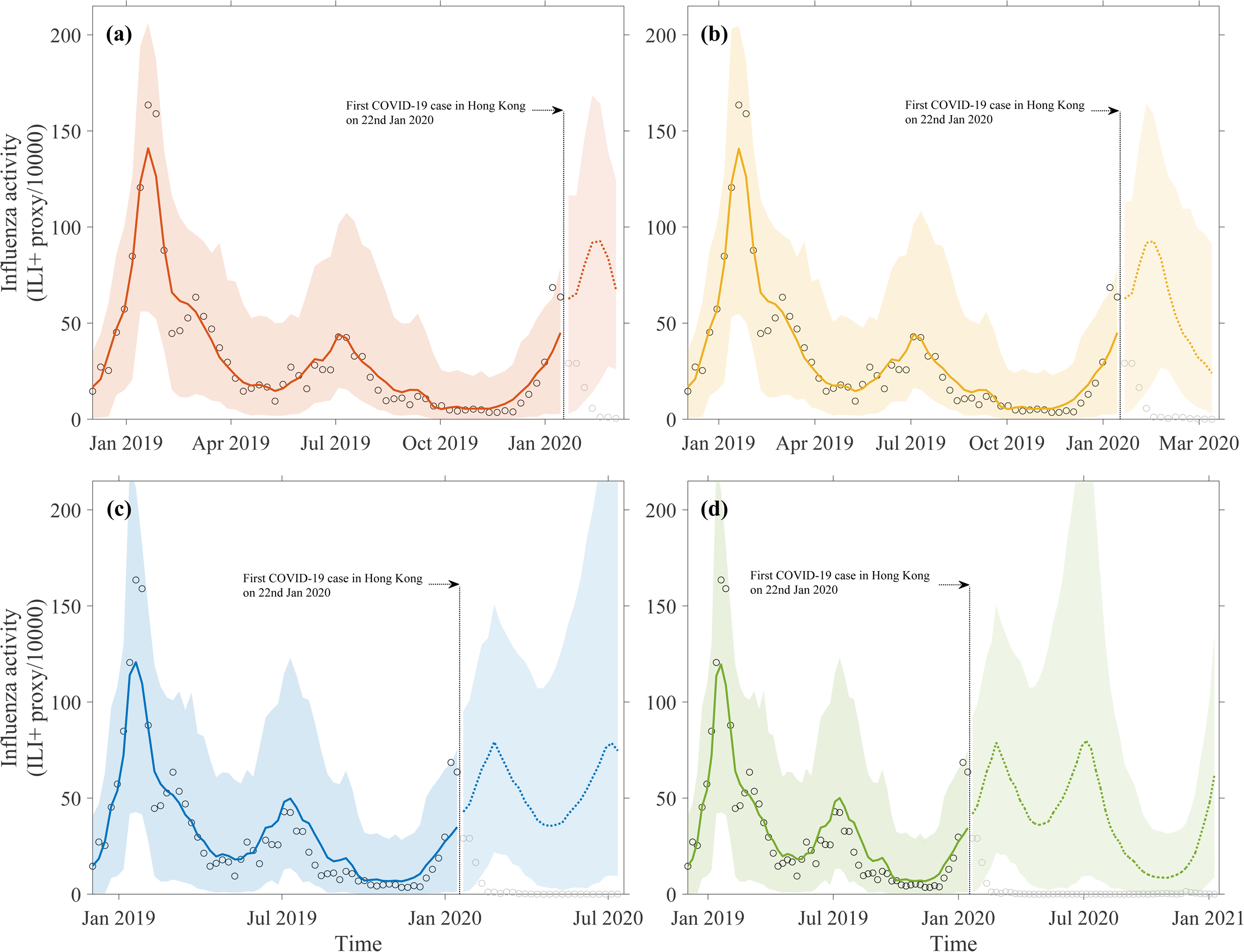
The forecast of influenza activity in Hong Kong starting from 22^nd^ January 2020, for different short-, medium and long-term forecast windows: (a) 7 weeks ahead, (b) 13 weeks ahead, (c) 26 weeks ahead, and (d) 52 weeks ahead. The black circles represent the observed influenza activities (weekly ILI+ proxy) with their predicted activity (fitted) in respective colored solid lines and the dashed lines represent the mean of ensemble forecast for different terms. The respective colored shades represent their corresponding 95% prediction intervals.

**Figure 5.**
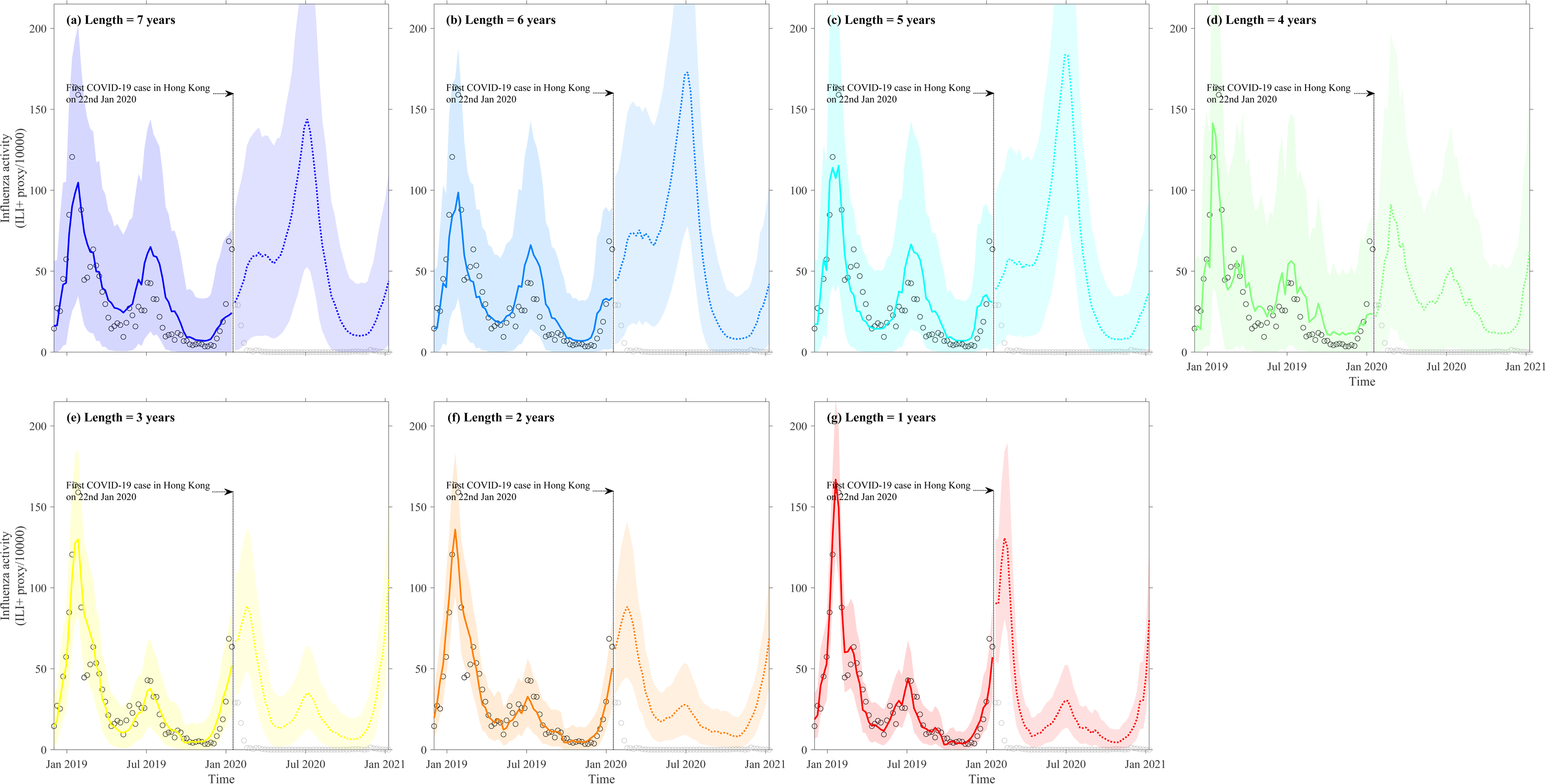
The counterfactual forecasting for 2020/21 season based on the optimal ensemble model by considering different lengths of historical data, (a)-(g) from 7 years to 1 years. The black circle denoted the observed influenza activity (ILI+ proxy/10^4^) and the solid lines in respective colors represent the model predictions with 95% prediction intervals in shades. The dashed lines represent the mean of forecasts 95% prediction intervals (in respective colored shades).

**Table 1.** Influenza forecasts for the 2019-20 winter season and successive seasons during COVID-19 pandemic for various forecasting outcomes, including peak activity, peak timing, attack rates, and associated reduction in attack rates due to the effects of PHSMs. The short, medium, and long-term forecasts were conducted since 22^nd^ January 2020 with 7, 13, 26, and 52 weeks ahead.

| **Forecasting Outcomes (95% prediction interval)** | **7 weeks ahead (22<sup>nd</sup> January 2020 to 11<sup>th</sup> March 2020)** | **13 weeks ahead (22<sup>nd</sup> January 2020 to 22<sup>nd</sup> April 2020)** | **26 weeks ahead (22<sup>nd</sup> January 2020 to 22<sup>nd</sup> July 2020)** | **52 weeks ahead (22<sup>nd</sup> January 2020 to 20<sup>th</sup> January 2021)** |
| --- | --- | --- | --- | --- |
| **Peak activity** | 94.8 (31.6,166.0) | 94.3 (30.3,167.7) | 1<sup>st</sup> peak: 75.0 (22.6, 127.0)<br>2<sup>nd</sup> peak: 96.9 (14.4,242.8) | 1<sup>st</sup> peak: 73.9 (22.4, 121.3)<br>2<sup>nd</sup> peak: 98.45 (14.4,243.4) |
| **Peak timing** | 23<sup>rd</sup> -29<sup>th</sup> Feb 2020 | 23<sup>rd</sup> -29<sup>th</sup> Feb 2020 | 1<sup>st</sup> peak: 1<sup>st</sup> -7<sup>th</sup> Mar 2020<br>2<sup>nd</sup> peak: 5<sup>th</sup> -11<sup>th</sup> Jul 2020 | 1<sup>st</sup> peak: 1<sup>st</sup> -7<sup>th</sup> Mar 2020<br>2<sup>nd</sup> peak: 5<sup>th</sup> -11<sup>th</sup> Jul 2020 |
| **Attack rates (%)** | 5.5 (1.5,9.2) | 8.0 (3.3,13.5) | 16.7 (6.0,36.5) | 23.7 (8.3, 53.0) |
| **Reduction in attack rates (%)** | 81.3 (45.4, 91.0) | 87.8 (73.9,93.7) | 93.5 (85.7,97.7) | 95.2 (89.1,98.3) |

### Counterfactual forecast and projection of influenza activity and its dynamical characteristics in post COVID-19 pandemic period since March 2023

Since 1^st^ March 2023, stringent PHSMs in Hong Kong were lifted, along with the cancellation of the mask mandate. Influenza activity gradually increased, reaching a peak (with 85 cases per 10,000 consultations) in mid-April. Subsequently, activity declined to lower levels between May and August, and a second peak occurred at the end of September (with a higher magnitude of 120 cases per 10,000 consultations). Further, in early 2024, activity continued to fluctuate, characterized by moderate peaks (with 65.83 cases per 10,000 consultations) in mid-January (**Fig. 1a**). Therefore, we observed a noticeable shift of seasonality of influenza transmission in post-pandemic period ^59^.

Theoretically, the reemerged epidemic should show a peak with a higher magnitude compared to the pre-pandemic period due to the high population susceptibility level ^46^. However, the cessation of these measures might not immediately eliminate their impact on the community ^56^. Moreover, influenza and SARS-CoV-2 display pronounced similarities in both their transmission dynamics and their cellular tropism for replication within the human respiratory tract ^57,58^. Therefore, we assumed two possible characteristics in the model to reconstruct the influenza transmission in post-pandemic period. (a) the effect of PHSMs would decrease linearly to zero at 0, 2, 4, 6, and 8 weeks following their cessation, under various residual stringency levels. (b) the various population susceptibility levels (accumulated during the lull period of 3 years) accounting for possible cross-competition (cross-protection) to influenza from COVID-19 infections^57,58^. We extended our framework for counterfactual forecasting and projection the outcomes in post-pandemic period under various reasonable parametric ranges of these characteristics (MATERIALS AND METHODS).

Following the cancellation of the mask mandate, the counterfactual post-pandemic forecasting and projection for the epidemics since 2023 under various combinations of reduced PHSM stringency and cross-protection impacts revealed that the scenario of a 70%-90% reduction in PHSM stringency and an 8%-14% reduction (over the baseline as on 31^st^ December 2022) in population susceptibility could mimic the observed outcomes better with lower root mean square errors (RMSEs), as shown in **Fig. 6**. The impact of PHSMs and cross-protection was found to be sensitive to forecasting and projection outcomes including peak timing, peak magnitude, and attack rate (**Fig. 7** and **Table S3).** For instance, under the extreme counterfactual scenario with 100% reduction in PHSMs impact and no cross-protection from COVID-19, it could lead up to an epidemic with 3-fold higher (over observed) peak magnitude in the week of 21^st^ – 27^th^ May 2023 (**Fig. 7**). Along with the combined impact, the individual impact of PHSM reduction and cross-protection effects found to be comparable (**Fig. S4)**. Furthermore, the sensitivity analyses on forecasting and projection start week following the cancellation of the mask mandate and underlying historical data lengths for model training, suggested a variation in the forecast and projection of outcomes (**Figs. S5–S8)**.

**Figure 6.**
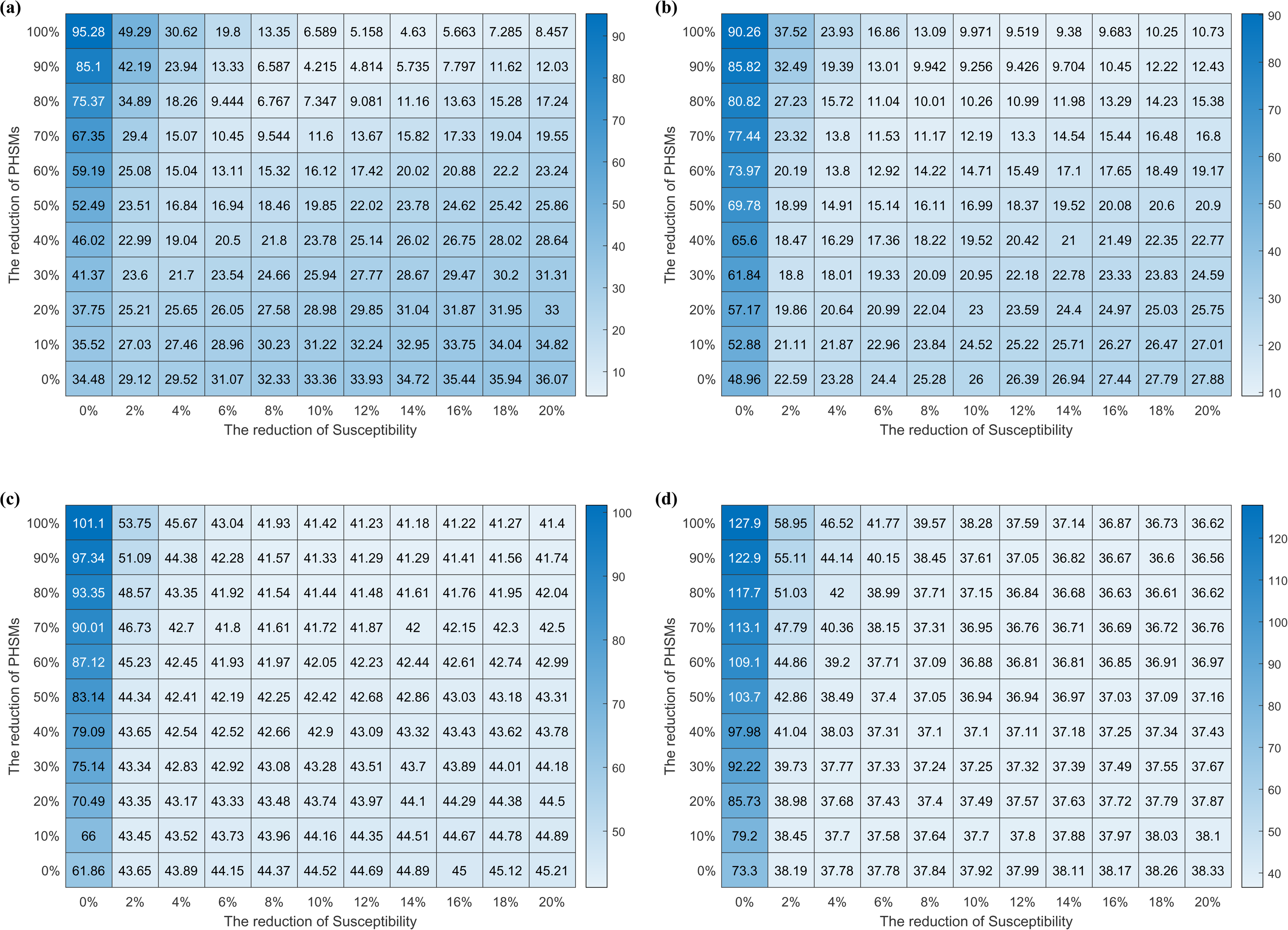
Counterfactual scenarios exploration for plausible reduction in susceptibility and PHSMs impact on the epidemics in post-pandemic period. The heatmap of the root mean square error (RMSE) between the counterfactual forecasting (projection) and the observed influenza activity in Hong Kong starting from 15^th^ April 2023, for different forecasting windows: (a) 7 weeks ahead, (b)13 weeks ahead, (c) 26 weeks ahead, and (d) 52 weeks ahead.

**Figure 7.**
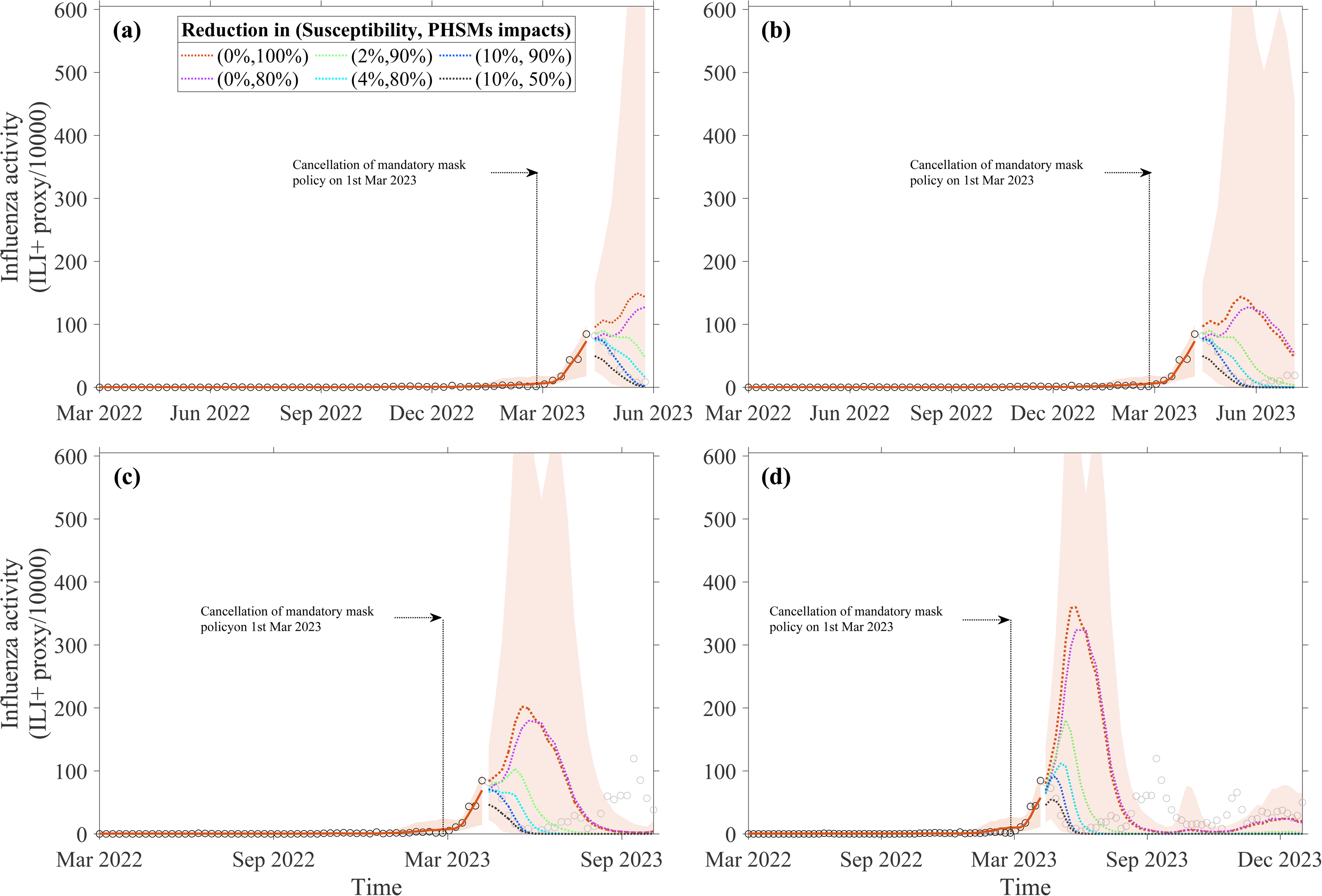
The forecast and projection of influenza activity starting from 15^th^ April, 2023, under various counterfactual scenarios for different short-, medium and long-term forecast windows: (a) 7 weeks ahead, (b) 13 weeks ahead, (c) 26 weeks ahead, and (d) 52 weeks ahead. The black circles represent the observed influenza activities (weekly ILI+ proxy) with their predicted activity (fitted) in respective colored solid lines and the dashed lines represent the mean of ensemble forecast for different terms and various plausible counterfactual scenarios. The respective colored shades represent their corresponding 95% prediction intervals. The baseline forecast scenarios (no reduction in accumulated susceptibility and complete cessation of PHSMs, i.e., (0%, 100%)) are presented by the dashed lines in the same color as the model fit. Forecasts for other combinations with plausible levels (reduction in susceptibility, PHSMs impact reduction) are shown in distinct respective colors, as specified in the legend.

## DISCUSSION

Influenza activity and its seasonality in a subtropical location, Hong Kong were found to be comparatively intricate with year-round circulation and multiple peaks (**Fig. 1a**), which was reported as the results of various potential climatic and social drivers ^10,22,37,46,47,60^. Further, the PHSMs for COVID-19 pandemic were reported to have a significant impact on influenza transmission and seasonality in Hong Kong during the pandemic (lull period) ^46,47,49^, and even in post-pandemic era by co-evoluting their inter-dynamics ^61–65^. We developed ensemble forecasting using multi-stream surveillance data on syndromic, virological, and extrinsic seasonal and social drivers at a temporal scale. Such ensemble models have often been suggested for better forecasting of respiratory viruses over an individual model ^13,43–45,66^. We employed temporal cross-validation to capture the temporal dynamics of the underlying forecasting outcomes and their potential drivers ^51–55^. We found that forecasting performances of different individual models varied significantly for different terms (short-term, medium-term and long-term) of forecasting and were sensitive to the length of historical data used (**Fig. 2**). We evaluated the forecasts of various outcomes using different lengths of historical data for pandemic period since January 2020 (**Fig. 5)** and post-pandemic period since March 2023 forecasts (**Fig. S9)**. The variation in forecasting outcomes (weeks ahead infections pattern, peak magnitude, and peak timing) suggested performing short-term (7 weeks ahead), medium-term (13 and 26 weeks ahead) and long-term (52 weeks ahead) forecasting individually with related adjustments in historical data length along with ensemble models^45^. Our analysis revealed that forecasts generated using historical training data of varying lengths can yield divergent outcomes. Relying on a single model or fixed dataset timeframe may introduce bias, thereby compromising the reliability and robustness of forecasting. To address this, we recommend employing ensemble models, which integrate multiple methodologies to enhance the reliability and robustness of forecasts. Furthermore, the variation in forecasting performance across short-, medium-, and long-term horizons (**Fig. 2 and 3**) suggests that forecast validation criteria should be applied separately to each forecast term rather than using a uniform criterion, ensuring more robust and reliable outcomes.

The counterfactual forecast for the 2019/20 winter season showed a significantly higher influenza activity with a peak in late February 2020, suggesting that COVID-19 PHSMs strongly mitigated influenza transmission in community. Several studies across the globe ^38,39,41,42^ reported similarly low levels of influenza activity during the 2019/20 winter season under the pandemic impact. However, impacts of COVID-19 PHSMs varied across countries and regions, and could be characterized by the prior influenza activities and stringency of the PHSMs in these locations^38^. In Hong Kong, the immediate impact of COVID-19 PHSMs was found in reducing the attack rate (up to 88% (74%, 94%)) for the 2019/20 winter influenza epidemic alone (**Fig. 4 and Table 1**). Similar reduction in attack rate for the 2019/20 winter season was reported for Hong Kong ^47^, United States ^40^, New Zealand ^41^, Australia ^67^, Korea ^41^, and mainland China ^40,68^; which have all shown a significant reduction in influenza activity during the COVID-19 pandemic. In the absence of effective pharmaceutical interventions, strategic implementation of these PHSMs would be a feasible and effective approach to prevent influenza epidemics ^40,49,68^.

For the counterfactual forecast and projection of post-pandemic season since March 2023, the framework could forecast the reemergence of influenza activity with a comparable peak magnitude following the relaxation of PHSMs (**Fig. 7**), likely due to the recovery of virus transmissibility and the increased population susceptibility resulting from the prolonged suppression of influenza activity in Hong Kong ^46^. However, the forecasted peak was not observed after the relaxation of PHSMs, possibly due to the combined effects of the residual impact of PHSMs in the population, coexistence/ circulation of influenza and COVID-19 with their potential cross-protection^39,63,69,70^, interactions with other respiratory diseases, changes in human behavior, and genetic changes influenza viruses ^71–73^. The counterfactual forecasting and projection during this post-pandemic period indicated a significant combined impact of PHSMs and cross-protection (reduced susceptibility) from the infection with other respiratory diseases including COVID-19 on influenza virus infections in Hong Kong (**Fig. 7, Table S3**).

The grid search on various possible scenarios characterized integrated mechanisms under plausible levels of PHSMs and population susceptibility in relation to observed influenza activity (**Fig. 6 and S4**). In particular, reduced impacts of PHSMs could increase the transmissions, but reduced population susceptibility could decrease the transmission, therefore their combined impacts could define the course of epidemics. The relationship between these factors and infections may not be a straightforward linear one, often characterized by non-linear impacts for community transmission ^46^. We found the scenarios with up to 10%-30% residual impact for PHSMs (over the baseline impact as on 15^th^ April 2023) in community after relaxation and 8%-14% reduction in population susceptibility (population immunity) could mimic better (lower RMSEs) the observed dynamics of influenza during post-pandemic period (**Fig. 7**). These findings are found to be much more evident with shorter term (7 and 13 weeks ahead) forecast, over longer term (26 and 52 weeks ahead) forecasts (**Fig. 7a and b**) which could be driven by longer historical impact. Studies reported on the impact of the PHSMs in community could not be immediately zero upon the relaxation in post-pandemic period ^56,59,74^, particularly for Hong Kong more than half of the residents (53.6%) continued wearing masks and avoiding various community activities even after (at least up to 3 weeks) cancelation of mandatory mask policy in March 2023 ^56,59^. Furthermore, the sensitivity analyses on forecasting start week suggested a significant variation on forecast of outcomes (**Figs. S5–S8)** presented counterfactual forecasts under different delays and stringency in the impacts of the cancellation of the mask mandate (Supplementary Section 3). On the other hand, in post-pandemic period, population susceptibility (immunity) could be effectively modulated by the lull period of 3 years, antigenic changes in the influenza viruses, and potential coexistence/cocirculation of other respiratory viruses including SARS-CoV-2 ^57,58^.

The ensemble forecasting framework was found to be equally compatible in performance for the influenza dynamics during pandemic and post-pandemic period with respective uncertainty (**Figs. 4 and 7**). While the forecasting and projecting the outcomes in post-pandemic period associated with significantly higher uncertainty compared to that of in pandemic period particularly for medium and long-term forecasts. These variations in the pattern and uncertainty could be characterized by the data lull period of 3 years with sporadic influenza circulation, hence significantly depending on the training window lengths, which could include the pre-pandemic data into consideration (**Fig. S9** and Supplementary Section 4). In particular, **Fig. S9(a–d**) suggested the forecasts that incorporated pre-pandemic data could predict a large peak with less uncertainty in the 2023/24 influenza epidemic following the relaxation of PHSMs. In contrast, the counterfactual forecasts of the outcomes with the data from the pandemic period only could have higher uncertainty and also failed to produce this significant peak (**Fig. S9(e–g**)). This discrepancy arose because the during pandemic period only, the models did not account for prior influenza activity, thereby overlooking the potential influenza immunity debt ^72,73^. These findings suggested that future influenza forecasts in the post-pandemic era should incorporate factors such as cross-protection and interactions between COVID-19 and other respiratory diseases ^71^.

The study has several limitations. First, we did not account for the evolution of the influenza virus over time, which could influence long-term transmission dynamics. Interactions between different virus strains might also create more complex transmission pathways ^75–77^. Second, our model did not incorporate demographic factors, such as population age, movements or variations in human behavior across the study period, which are closely associated with influenza susceptibility and transmissibility^78^. Including these factors could improve the accuracy of influenza activity forecasts, although we used an ensemble approach to enhance forecasting performance. Third, we considered temporal cross-validation with a sliding window of 7 weeks for short-, medium- and long-term forecasts for computational efficiency, which could limit the forecasting outcome for shorter terms with less than 7 weeks (1 to 6 weeks) ahead. While ensemble models generally require more computational resources than individual models, our methodology supports parallel training to manage these demands. Fourth, we did not have the scope to validate the forecast performance for post-pandemic period under the lull period of influenza activity, which might result in a comparably inferior forecast with higher uncertainty in post-pandemic period (**Fig. 7**), despite that ensemble model is less sensitive to the data as long training window is also assimilated. Finally, we assumed the detection rate of influenza remained constant throughout the study period. However, this assumption might not hold true, especially since a significant portion of the detection effect has been allocated to COVID-19 during the pandemic and the surveillance capacity for influenza detection might not have been immediately following the relaxation of PHSMs in March 2023.

In conclusion, we could forecast influenza activity and assess the dynamical characteristics during and post-pandemic periods in Hong Kong, a (sub-)tropical city, by developing multi-stream data-driven ensemble models. We observed that the model performance and forecasting of outcomes could vary with the length of historical data and the forecasting windows. The forecasting for 2019/20 winter season indicated the significant role of COVID-19 related PHSMs on influenza activities. The influenza forecasting and projection for 2023/24 season suggested that the change in transmission dynamics and seasonality of influenza viruses in post-COVID-19 era, emphasizing the additional factors including population immunity under the co-circulation with COVID-19 should be considered in future influenza forecasts. Our proposed forecast framework, extendable to other respiratory diseases and locations, can offer healthcare agencies predictive data on future infection rates, thereby enhancing preparedness.

## MATERIALS AND METHODS

### Time series of influenza activity (ILI+ proxy)

We retrieved syndromic and virological data including weekly numbers of confirmed influenza cases, total specimens tested, patient visits for influenza-like illness (ILI), and total consultations during January 2010 to August 2024 from the sentinel and laboratory-based surveillance networks and Centre for Health Protection of the Hong Kong Special Administrative Region. We derived ILI+ proxy a measure of influenza virus activity in the community, calculated by multiplying the weekly ILI rates and the influenza-positive rates and standardized for the sentinel sites in the surveillance network (**Fig. 1a**). Such proxy was found to have better linear correlation for influenza virus activity in community^4,46,47^.

### Time series of meteorological and environmental drivers, school closures and COVID-19-related PHSMs

Previous studies identified ambient absolute humidity, temperature, and ozone concentration as significant drivers of influenza in Hong Kong ^31^. We obtained daily mean relative humidity, temperature, and ozone concentration index data from the Hong Kong Observatory and the Hong Kong Environmental Protection Department during the study period (**Fig. 1b-d**). We calculated the mean absolute humidity using relative humidity and temperature ^48^. School closures and holidays were also reported as potential factors in reducing influenza transmissibility in Hong Kong and other regions ^32–34^. We retrieved the information on scheduled holiday-related school closures (e.g., Chinese New Year, winter holidays, summer holidays, and public holidays) as well as reactive school closures during seasonal epidemics and pandemics in Hong Kong. Approximately 95% of children in Hong Kong attended local government schools (**Fig. 1e**).

The recent COVID-19 pandemic and associated PHSMs were reported to have a significant impact on influenza activity in communities across the globe ^40,46,49^. We obtained data on these measures for Hong Kong from the Oxford COVID-19 Government Response Tracker (OxCGRT) ^50^. The OxCGRT compiled information on the timing and stringency of various government policies, which were used to generate a set of policy indicators, as illustrated in **Fig. 1e**.

### Mechanistic model construction

We applied state-space models of the susceptible-vaccinated-exposed-infectious-recovered-susceptible (SVEIRS) type, incorporating a time-varying transmission rate, *β*(*t*), driven by the time series of various drivers. A schematic illustration of the mechanistic model with associated model parameters is provided in **Fig. S1** and **Table S1**. To capture less regular influenza seasons with annual and biannual peaks for Hong Kong, a baseline model was constructed assuming that the transmission rate followed a Fourier series (Supplementary Section 1). Further, the baseline model was improved by incorporating the effects of significant extrinsic drivers on transmissibility under a multiplicative framework. For absolute humidity, the U-shaped forms of association with influenza transmission were considered ^26,36,37^. Temperature was included with a non-linear negative association with transmission ^27^, was adjusted for average levels and ambient ozone concentrations were included as a negative form of association with transmissibility ^29–31^. The effects of school closures and holidays were included as the categorical variable on their timing to account for the impact on transmission dynamics ^32,33,35,34^. During the pandemic period (from January 2020 to February 2023) we considered the impact of PHSMs on transmission ^38–42^, by using their implication timing and stringency index across the study period. We considered a series of predictive model variants (16 individual models) by exploring various combinations of these drivers, and different lengths of training periods with historical data (Table S2).

### Likelihood and estimation of model parameters

The parameters of the predictive models (listed in Table S1) were estimated using a Markov chain Monte Carlo (MCMC) method (Supplementary section 5). By considering those transferred from the exposure to infectious compartments, the modelled influenza incidence number at day *t* detected in the surveillance system was 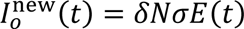, where *E*(*t*) denoted the proportion of exposed individuals, 1/σ denoted mean latent period and *δ* denoted the ascertainment rates. Thus, the modelled weekly influenza activity at week *w* was the cumulative sum of modelled daily incidence, i.e. 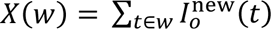. We assumed the square root of the observed influenza activity at week *w* 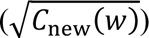 followed a normal distribution, with a mean equal to the square root of the modelled weekly influenza activity at week *w* 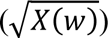 and a variance of *ϵ*^2^.

### Weighting of model variants for ensemble forecasting

Ensemble forecasting provides more robust and less biased forecasting compared to the individual models ^10,43–45^. To evaluate the performance of the ensemble models, we ranked the model variants using the cross-validated weighted interval score (WIS) ^51^ as stated in the sub-section below. Individual forecasts were then aggregated to produce the final ensemble forecast using a rank-order centroid (ROC) weighting approach, which demonstrated superior accuracy compared to other rank-based weighting schemes ^52^. The ROC weight was calculated based on the ranks of individual models contributing to the ensemble model. Specifically, the weight (*ω*_*j*_) assigned to a model with rank *j* of *n* was defined as follows:

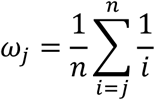

We constructed 5 ensemble models for forecasting using 16 individual models under various combinations of significant drivers including baseline model, which were trained on 7 different lengths (1-7 years) of historical data (details are in Table S2). The best ensemble model was chosen based on the forecasting performance of its individual models and utilized for short-term (up to 7 weeks ahead), medium-term (up to 13-26 weeks ahead), and long-term (beyond 26 weeks) forecasting of the outcomes.

### Temporal cross-validation and assessment of forecast performance

We employed a temporal cross-validation method ^53–55^ to evaluate the forecasting performance of the predictive models, which were trained on influenza activity and multi-stream surveillance data with temporal dependencies. The forecasting performance of all 16 individual model variants and 5 ensemble models was then compared. The temporal cross-validation approach divided the time series into sequential training and validation sets. Each training set spanned a fixed time window, followed by *M* observations in the validation set, where *M* represented the forecast window length in week. Forecasting performance was assessed using a rolling forecasting origin across the data series. To evaluate different terms of forecast performance, we conducted temporal cross-validation with *M* ∈ {7, 13,26,52}. Additionally, we tested the impact of the training window size on performance by using seven different training window sizes with 1-7 years. Further details on the temporal cross-validation method are provided in Supplementary section 6.

We evaluated the forecast performance of our proposed models using the weighted interval score (WIS) ^51^, a metric designed for assessing epidemic forecasts in an interval format. Unlike methods that focus solely on single-point estimates, WIS accounts for penalties when prediction intervals fail to include the observed value, thus providing a measure of how well the model captures uncertainty. WIS generalizes the concept of absolute error to probabilistic forecasts, decomposing performance into measures of accuracy and penalties for over-or under-prediction. A lower WIS score indicates better forecasting performance.

### Forecasting outcomes of influenza activity and assessing their dynamical characteristics during and post-COVID-19 pandemic periods

We defined various forecasting outcomes of influenza activity, including peak timing, peak intensity, and attack rate under 4 different time horizons of short-term, medium-term, and long-term forecasting schemes. We used the posterior parameter samples, obtained from the MCMC evaluation to forecast influenza activity outcomes in Hong Kong since 22^nd^ January 2020 (the first COVID-19 case reported) to assess the seasons during the pandemic period. For the post-pandemic scenario, the cancelation of the mandatory mask policy was lifted on 1^st^ March 2023, but it was unlikely to be disappeared immediately in the community, and thus we considered different delays (0, 2, 4, 6, and 8 weeks) since 1^st^ March to assess the seasons in the post-pandemic period ^56^. Despite forecasting 2019/20 winter season and successive seasons were considered to assess the impact of COVID-19 PHSMs on the influenza activity during the pandemic, the 2023 spring season and successive seasons for assessing the post-pandemic variation in dynamical characteristics of influenza in the community.

During COVID-19 pandemic period, we compared the observed influenza activity with the retrospective forecast outcomes by calculating the reduction in activity by comparing observed influenza activity (*C*_new_(*w*)) and forecasted influenza activity X(*w*) at week *w*. The forecasted attack rates in 10000 consultations over a given time interval were defined as *AR*_*F*_ = ∑_*w*_ X(*w*)⁄10000, and the observed attack rates were defined as *AR*_*o*_ = ∑_*w*_ *C*_*new*_(*w*)⁄10000, hence the reduction rate in attack rates was derived as 1 − *AR*_*o*_/*AR*_*F*_^69^. For post-pandemic period, community activity of influenza reemerged since 2023 spring season after the relaxation of PHSMs including the cancellation of face-mask mandate in Hong Kong. Looking at the shift of seasonality of influenza transmission, we hypothesized that along with extrinsic drivers, two additional key factors could drive such shift in post-pandemic period. First, there were still residual impact of COVID-19-related PHSMs in the community and population behavior even after their relaxation or cancellation ^56^. Second, since both influenza and SARS-CoV-2 exhibit considerable similarities in their modes of transmission and their cellular tropism, the population susceptibility could be reshaped by possible cross-reactive immunity (cross-competition) to influenza from repetitive COVID-19 infections and vaccinations and genetic changes in the viruses ^57,58^. We extended this framework for forecasting the post-pandemic period under various reasonable ranges of these two parameters. We explored counterfactual forecasting under different reduction levels of PHSMs impact and population susceptibility. We explored a wide range of reduction levels of PHSMs as (0% −100%) over the suitable baseline stringency of PHSMs as on 31^st^ December 2022, as *P*(*t*) = (1 − *r*_*p*_) ∗ *P*_0_, where *r*_*p*_ = *t/D*_*t*_ denoted the reduction rate (%), *D*_*t*_ was the delayed time from 1^st^ March 2023, and *P*_0_ = 11.11 denoted the value of PHSM stringency index as of 31^st^ December 2022, which was assumed to be unchanged until 1^st^ March 2023 (**Fig 1e**). We considered a possible range of population susceptibility reduction (0% to 20%) at a time-varying scale with COVID-19 infection proxy trajectory by incorporating a multiplicative component as 1− *r*_*c*_ ∗ COVID(*t*)), i.e, *S*(t) = *S*(*t*)*(1− *r*_*c*_ ∗ COVID(*t*)), where COVID(*t*) denoted the COVID-19 infection proxy during the forecasting period, and *r*_*c*_ denoted reduction in population susceptibility to influenza due to the impact of cross-protection/competition of COVID-19 on influenza. Finally, we identified the best possible estimates of these two parameters in mimicking the observed influenza activity in post-pandemic period by comparing the RMSE (root mean square error) measure.

## Data Availability

Data deposition: The data and code used in this analysis will be made publicly available and deposited on GitHub, https://github.com/WangDongHKU/Ensemble_Influenza_Forecasting.

## ACKNOWLEDGEMENTS

The authors thank Julie Au for technical assistance. This project was supported by the grants from the Health and Medical Research Fund (project no. 18171202); AIR@InnoHK administered by Innovation and Technology Commission, Government of the Hong Kong Special Administrative Region. The funding bodies had no role in study design, data collection and analysis, preparation of the manuscript, or the decision to publish.

## Contributors

All authors meet the International Committee of Medical Journal Editors criteria for authorship. STA, DW, and BJC conceived the study and designed the statistical and modelling methods; DW, YCL, SS, and STA collected, assimilated, and analyzed the data; and DW and STA wrote the first draft of the manuscript. All authors contributed to the interpretation of the results, revised the manuscript critically for intellectual content, and gave final approval of the version to be published.

## Supplementary Material

Supplementary information

