## SUPPLEMENTARY MATERIALS for "Ensemble forecasting of influenza activity and assessing its year-round dynamical characteristics during and post-COVID-19 pandemic periods in a sub-tropical location"

* Communicating author

† These authors contributed equally

### 1. State-space mechanistic transmission models formulation and their ensembling for influenza forecasting

In this study, we employed a susceptible-vaccination-exposed-infectious-recovered-susceptible (SVEIRS) framework to model influenza transmission dynamics in the (sub-)tropical city of Hong Kong. The model incorporates multi-stream surveillance data, including absolute humidity, temperature, ozone levels, and information on school closures and holidays.

The structure of our model is as follows:

$$\frac{dS(t)}{dt} = \mu + q_1R(t) + q_2V(t) - \beta(t)S(t)I(t)^m - (\mu + v(t))S(t)$$
$$\frac{dV(t)}{dt} = v(t)S(t) - (1-x)\beta(t)V(t)I(t)^m - (\mu + q_2)V(t)$$
$$\frac{dE(t)}{dt} = \beta(t)I(t)^m(S(t) + (1-x)V(t)) - (\sigma + \mu)E(t)$$
$$\frac{dI(t)}{dt} = \sigma E(t) - (\gamma + \mu)I(t) + \alpha$$
$$\frac{dR(t)}{dt} = \gamma I(t) - (q_1 + \mu)R(t)$$

Where $S(t), E(t), V(t), I(t)$, and $R(t)$ denoted the proportion of susceptible individuals, exposed individuals, vaccinated individuals, observed infectious individuals, unobserved infectious individuals, and recovered individuals at day t, respectively. $\mu$ was the birth/death rate was set to be 0.00918 per person-year [2,3]. $\beta(t)$ was the time-varying transmission rate. An infected individual would become infectious after $1/\sigma$ days on average since infection (i.e. mean latent period), which was assumed to be 1.5 days [4,5]. The individual would be infectious for $1/\gamma$ days on average (i.e. mean infectious period), which was assumed to be 5.5 days [5-8], and be recovered afterward. The immunity in the recovered individual would wane over time and protect the individual for $1/q_1$ days on average, which was assumed to be 450 days [7,9,10], and the immunity from vaccination would protect the individual for $1/q_2$ days, which was assumed to be 270 days [11,12]. $\nu(\mathrm{t})$ and $x$ denoted the seasonal vaccination rate from October to December each year and the vaccine efficacy, respectively, which were set to 17% and 59% [12-15]; whereas $\nu(t) = 0$ in other months. Besides, $m = 0.97$ denoted the heterogeneous population mixture [2,16]. $\alpha$ represented seeding from outside the model population [2]. A schematic illustration of the mechanistic model is provided in **Fig. S1**, while **Table S1** details the corresponding model parameters, along with the sources of initial values and prior distributions.

Considering annual and bi-annual influenza waves in Hong Kong, we considered a baseline transmission rate that follows a Fourier series as,

$$\beta_{base}(t) = exp\left(a_0 + a_1\, cos\left(\frac{4\pi t}{365.25}\right) + a_2\, sin\left(\frac{4\pi t}{365.25}\right) + a_3\, cos\left(\frac{2\pi t}{365.25}\right) + a_4\, sin\left(\frac{2\pi t}{365.25}\right)\right),$$

where $a_0, a_1, a_2, a_3, a_4$ indicated the coefficient of the baseline seasonality of transmission rate.

Given the baseline transmission rate, we accounted for the impact of the abovementioned drivers on influenza transmission in Hong Kong under multiplicative assumption. Assuming the U-shape association for absolute humidity with influenza transmission [2,17], we defined humidity-associated components as $\beta_{AH}(t) = exp([b_1 q^2(t) + b_2 q(t)])$, where $q(t)$ denoted the mean absolute humidity at time point t. $\mathrm{b}_1$, $\mathrm{b}_2$ were the coefficients for the response of absolute humidity on influenza transmission rates. Several studies reported a non-linear negative association of a mean ambient temperature on influenza transmission [2,18,19]. Therefore, we considered the temperature-driven component as $\beta_T(t) = exp\,(-cT(t))$, where $T(t)$ was the mean temperature at time $t$, and $c$ was the coefficient, indicating the response in transmission rate for associated temperature changes. While ambient ozone concentrations were reported to have a negative association with influenza transmissibility [20,21], hence we defined the ozone-driven component as $\beta_O(t) = exp\,(-dO(t))$, where $O(t)$ was the ambient mean ozone concentration at time t, and d was the coefficient, indicating the response in transmission rate for given ozone changes. Further, we introduced the effect of school closures/holidays on influenza transmission and defined the associated component as $\beta_H(t) = exp\big(-\xi H(t)\big)$, where $H(t)$ denoted the holiday data at time t, and ξ was the coefficient for the impact of school closures and holidays on the transmission rate. For the fitting from January 2020 to Feb 2023 when PHSMs were implemented, we considered the impact of PHSMs. We introduced a coefficient $\kappa \in (0,1]$ to describe the impact of PHSMs, i.e., $exp(-\kappa \cdot P(t))\beta(t)$ where $P(t)$ is the PHSMs index time series [22,23].

In our analysis of time-varying transmission rates, we evaluated all possible combinations of contributing factors, resulting in 16 individual models, as detailed in **Table S2**. To enhance the reliability of predictions, we adopted an ensemble forecasting approach that integrates these individual models with different lengths of historical data training windows, following established methodologies[24,25]. Five ensemble forecasting models were developed, each incorporating all factors except the baseline model, which excluded any factors. Specifically:

Ensemble Model 0 (E-0): Aggregated the zero-factor individual models

Ensemble Model I (E-I): Aggregated the one-factor individual models

Ensemble Model II (E-II): Aggregated the two-factor individual models

Ensemble Model III (E-III): Aggregated the three-factor individual models

Ensemble Model IV (E-IV): Aggregated the four-factor individual models

#### 2. Sensitivity analysis of starting points in forecasting since January 2020

We conducted a sensitivity analysis to examine the impact of different starting points on our forecasts. For the 2020/21 forecast, two alternative start dates were considered: 6th January 2020, when screenings were initiated at airports and train stations with connections to Wuhan, and 15th January 2020, the week preceding the first confirmed COVID-19 case in Hong Kong on 22nd January 2020. The forecasting results for these starting points across various terms were shown in **Figs. S2-3**. Our analysis revealed that the choice of starting point has only a minimal effect on the forecasting of influenza activities.

#### 3. Sensitivity analysis of starting points in forecasting since March 2023

The last PHSM (e.g., mask mandate) was lifted on 1st March, 2024. We tested the impact of PHSMs relaxation and viral competition with COVID-19 on influenza activity using counterfactual forecasting. This analysis assumed a 6-week delay in PHSM relaxation (**Fig. S4**). The results align with our intuitions: greater relaxation of PHSMs correlated with higher influenza transmission, while stronger viral competition with COVID-19 correlated with reduced influenza activity. Moreover, for the 2023/24 forecast in the post-pandemic era, we considered four additional forecasting starting points: 1st March, 15th March, 29th March, and 26th April, corresponding to 0, 2, 4, and 8 weeks after the lifting of the mandate. It was assumed that PHSMs would decrease linearly to zero from 1st March until each respective starting point, after which the forecasting process commenced. The results of these forecasts are presented in **Figs. S5–8**.

Compared to the 2020/21 forecasts, the 2023/24 forecasts exhibited notable sensitivity to the starting points. Taking long-term forecasting as an example, the forecast starting on 1$^{st}$ March predicted a smaller peak during the week of 11$^{th}$ –17$^{th}$ June, 2023, with a magnitude of 199.19 (0.75–549) (**Fig. S5(d)**). This forecast also showed a huge winter peak during the week of 24$^{th}$ –30$^{th}$ December, 2023. Similar forecasting pattern for the starting point from 15$^{th}$ March 2023 (**Fig. S8(d)**). In contrast, the forecast starting on 1$^{st}$ April and 26$^{th}$ April predicted a larger peak at 21$^{st}$ –27$^{th}$ May, 2023, with a mean magnitude of over 300, (**Figs. S7(d), S8(d)**), but no distinct winter peak was observed.

#### 4. Sensitivity analysis of different lengths of historical data in forecasting

**Fig. S9** illustrates the impact of historical data lengths on forecasting influenza activity since April 15, 2023. Panels (a) to (g) demonstrate how different time spans (ranging from seven to one year) affect forecasted influenza trends. Historical datasets spanning 4–7 years, which include pre-pandemic periods, effectively capture long-term seasonal patterns and variations in susceptibility, forecasting a larger peak in mid-May 2023. In contrast, datasets limited to the COVID-19 pandemic period primarily reflect recent epidemic characteristics but fail to account for long-term seasonality. Notably, forecasts derived from three-year historical datasets are more influenced by the prolonged epidemiological effects of the COVID-19 pandemic than those using 1–2 year datasets and do not forecast the peak in October 2023. Given that models trained on different lengths of historical datasets produce divergent forecasts and may introduce bias, we recommend ensemble forecasting to integrate insights from multiple data timescales, thereby improving overall reliability.

#### 5. Parameter estimation and model fitting

The mechanistic model was resolved using the Euler method, with a day as the time step for various model parameters (**Table S1**). Model fitting employed the adaptive Metropolis-Hastings algorithm to perform Markov Chain Monte Carlo (MCMC) sampling, implemented via the "mcmcstat" package in MATLAB 2020a (https://mjlaine.github.io/mcmcstat) [26]. Four chains were run, each consisting of 1,000,000 iterations, with the first 500,000 iterations discarded as burn-in. Convergence of the posterior distribution was evaluated using the Gelman-Rubin diagnostic.

#### 6. Temporal cross-validation

In this study, we employed a temporal cross-validation (CV) approach to select the optimal model for forecasting [27-29]. The dataset was divided into 70% for training and 30% for validation. Influenza activity data from January 2010 to December 2016 served as the training set, while data from January 2017 to December 2019 was used for validation. Using a 7-year training window as an example, the model was initially trained with data from the first week of January 2010 to the last week of December 2016. Validation was performed using data from week 1 to week $M$ in 2017, where $M$ represents the length of the forecasting window. In the subsequent step, the training window was rolled forward by 7 weeks (i.e., training data from week 8 of 2010 to week 7 of 2017), and validation was conducted for data from week 8 to 7+ $M$ in 2017. This rolling origin method was repeated until validation reached the last week of 2016. For other lengths of training periods from 6 to 1 year, the start points of training windows were 1$^{st}$ week of 2011, …, 1$^{st}$ week of 2016, respectively. All temporal cross-validation iterations followed a consistent process and shared the same validation periods of data. Forecasting performance was evaluated across different forecasting terms by varying the forecasting period $M$ (i.e., $M \in 7, 13, 26, 52$).

### 7. Supplementary tables

**Table S1. Descriptions of model parameters and their estimations for reliable choice of prior distributions.**

| Parameters | Description | Value/Prior distribution | References |
|---|---|---|---|
| $\mathbf{1/\sigma}$ | The mean latent period of the infected individuals (days) | 1.5 | [4,5,30] |
| $\mathbf{1/\gamma}$ | The infectious period of infected individuals (days) | 5.5 | [5-8,30] |
| $\boldsymbol{\mu}$ | The death rate (year-1) | 0.00918 | [2,3] |
| $\mathbf{1/q_1}$ | The duration of influenza immunity from infection (days) | 450 | [7,10,31,32] |
| $\mathbf{1/q_2}$ | The duration of influenza immunity from vaccination (days) | 270 | [12,33] |
| $\boldsymbol{v(t)}$ | The winter seasonal vaccination rate from October to December | 17% | [12,13] |
| $\boldsymbol{x}$ | The vaccine efficacy | 0.59 | [13-15] |
| $\boldsymbol{m}$ | An exponent to denote heterogeneous mixing | 0.97 | [34] |
| $\boldsymbol{\alpha}$ | random seeding from outside the model population | Uniform$(0, 0.01)$ | Model estimates |
| $\boldsymbol{a_0}$ | A coefficient determines the strength of the impact of seasonality. | Uniform $(-10, 10)$ | Model estimates |
| $\boldsymbol{a_1}$ | A coefficient determines the strength of the impact of seasonality. | Uniform $(-10, 10)$ | Model estimates |
| $\boldsymbol{a_2}$ | A coefficient determines the strength of the impact of seasonality. | Uniform$(-10, 10)$ | Model estimates |
| $\boldsymbol{a_3}$ | A coefficient determines the strength of the impact of seasonality. | Uniform $(-10, 10)$ | Model estimates |
| $\boldsymbol{a_4}$ | A coefficient determines the strength of the impact of seasonality. | Uniform $(-10, 10)$ | Model estimates |
| $\boldsymbol{\delta}$ | The ascertainment rates of infected individuals | Uniform $(0, 1)$ | Model estimates |
| $\boldsymbol{b_1}$ | A coefficient determining the strength of the impact of absolute humidity | Uniform $(0, 1)$ | Model estimates |
| $\boldsymbol{b_2}$ | A coefficient determining the strength of the impact of absolute humidity | Uniform $(-1, 0)$ | Model estimates |
| $\boldsymbol{c}$ | A coefficient determining the strength of the impact of temperature | Uniform $(0, 3)$ | Model estimates |
| $\boldsymbol{d}$ | A coefficient determining the strength of the impact of ozone | Uniform $(0, 1)$ | Model estimates |
| $\boldsymbol{\xi}$ | A coefficient determining the strength of the school-term impact | Uniform $(-1, 1)$ | Model estimates |
| $\boldsymbol{S(0)}$ | The initial proportion of susceptible population | Uniform $(0, 1)$ | Model estimates |
| $\boldsymbol{I(0)}$ | The initial proportion of unobserved infected population | Uniform $(0, 1)$ | Model estimates |
| $\boldsymbol{E(0)}$ | The initial proportion of exposed population | Uniform $(0, 1)$ | Model estimates |
| $\boldsymbol{R(0)}$ | The initial proportion of removed population | Uniform $(0, 1)$ | Model estimates |

| | | | |
|---|---|---|---|
| $\boldsymbol{V(0)}$ | The initial proportion of vaccinated population | Uniform $(0, 1)$ | Model estimates |
| $\boldsymbol{k}$ | The coefficient of PHSM effects | Uniform $(0, 1)$ | Model estimates |

**Table S2. The proposed five ensemble models comprise their associated individual model variations and corresponding combinations of factors.**

| **Ensemble models** | **Individual model variants** | **Extrinsic drivers included** | **Time-varying transmission rates ($\boldsymbol{\beta}(\mathbf{t})$ =)** |
|---|---|---|---|
| **E-0** | Baseline (B) | NA | $\beta_{base}(t)$ |
| **E-I** | B+AH | Absolute humidity (AH) | $\beta_{base}(t)\beta_{AH}(t)$ |
| | B+T | Temperature (T) | $\beta_{base}(t)\beta_{T}(t)$ |
| | B+O | Ozone (O) | $\beta_{base}(t)\beta_{O}(t)$ |
| | B+H | Holidays (H) | $\beta_{base}(t)\beta_{H}(t)$ |
| **E-II** | B+AH+T | AH, Temperature | $\beta_{base}(t)\beta_{AH}(t)\beta_{T}(t)$ |
| | B+AH+O | AH, Ozone | $\beta_{base}(t)\beta_{AH}(t)\beta_{O}(t)$ |
| | B+AH+H | AH, Holidays | $\beta_{base}(t)\beta_{T}(t)\beta_{O}(t)$ |
| | B+T+O | Temperature, Ozone | $\beta_{base}(t)\beta_{T}(t)\beta_{H}(t)$ |
| | B+T+H | Temperature, Holidays | $\beta_{base}(t)\beta_{T}(t)\beta_{H}(t)$ |
| | B+O+H | Ozone, Holidays | $\beta_{base}(t)\beta_{O}(t)\beta_{H}(t)$ |
| **E-III** | B+AH+T+O | AH, Temperature, Ozone | $\beta_{base}(t)\beta_{AH}(t)\beta_{T}(t)\beta_{O}(t)$ |
| | B+AH+T+H | AH, Temperature, Holidays | $\beta_{base}(t)\beta_{AH}(t)\beta_{T}(t)\beta_{H}(t)$ |
| | B+AH+O+H | AH, Ozone, Holidays | $\beta_{base}(t)\beta_{AH}(t)\beta_{O}(t)\beta_{H}(t)$ |
| | B+T+O+H | Temperature, Ozone, Holidays | $\beta_{base}(t)\beta_{T}(t)\beta_{O}(t)\beta_{H}(t)$ |
| **E-IV** | B+AH+T+O+H | AH, Temperature, Ozone, Holidays | $\beta_{base}(t)\beta_{AH}(t)\beta_{T}(t)\beta_{O}(t)\beta_{H}(t)$ |

**Table S3. Influenza forecasts for 2023 spring season and successive seasons in post COVID-19 pandemic for various forecasting outcomes, including peak activity, peak timing, attack rates, and associated reduction in attack rates due to the effects of PHSMs and cross-protection of COVID-19 on influenza.** The short, medium and long-term forecasts were conducted since 15th April 2023 with 7, 13, 26 and 52 weeks ahead respectively, under various levels of residual impact of PHSMs and susceptibility.

| **Forecasting Outcomes (95% prediction interval)** | **Reduction of (Susceptibility, PHSMs, )** | **7 weeks ahead (15th April 2023 to $3^{rd}$ June 2023)** | **13 weeks ahead (15th April 2023 to $15^{th}$ July 2023)** | **26 weeks ahead (15th April 2023 to $14^{th}$ October 2023)** | **52 weeks ahead (15th April 2023 to $13^{th}$ April 2024)** |
|---|---|---|---|---|---|
| **Attack rates (%)** | (0, 100%) | 8.3 (0.7, 30.8) | 13.5 (2.1, 47.8) | 19.6 (0.7, 57.0) | 36.9 (2.7, 62.1) |
| | (0%, 80%) | 6.7 (0.7, 24.4) | 12.3 (0.7, 45.1) | 18.6 (0.7,50.9) | 35.2 (2.9, 57.8) |
| | (2%,90%) | 5.1 (0.6,18.0) | 5.9 (0.6, 22.0) | 7.2 (0.59, 23.4) | 11.7 (1.5, 24.7) |
| | (4%,80%) | 3.4 (0.5,11.2) | 3.6 (0.6,12.9) | 4.1 (0.53, 12.3) | 6.2(1.2, 12.8) |
| | (10%,90%) | 2.6 (0.4, 7.1) | 2.6 (0.4, 7.2) | 2.7 (0.42, 7.4) | 3.8 (0.9, 7.8) |
| | (10%,50%) | 1.5 (0.4, 3.9) | 1.5 (0.40, 4.1) | 1.6 (0.39, 4.1) | 2.3 (0.6, 4.4) |
| **Peak activity** | (0, 100%) | 144.5 (0.09,716.9) | 145.5 (0.04,720.4) | 202.1 (0.01, 708.0) | 358.6 (0.4,716.1) |
| | (0%, 80%) | 126.4 (0.03,617.3) | 122.9 (0.01, 582.8) | 178.5 (0.01, 613.7) | 322.3 (0.1,619.2) |
| | (2%,90%) | 84.63 | 84.63 | 103.4 (0.03,394.5) | 162.3 (0.04,377.1) |
| | (4%,80%) | 84.63 | 84.63 | 84.63 | 106.1 (0.1,239.5) |
| | (10%,90%) | 84.63 | 84.63 | 84.63 | 84.63 |
| | (10%,50%) | 84.63 | 84.63 | 84.63 | 84.63 |
| **Peak timing** | (0, 100%) | $21^{th}$ – $27^{th}$ May 2023 | $21^{th}$ – $27^{th}$ May | $21^{th}$ – $27^{th}$ May | $21^{th}$ – $27^{th}$ May |
| | (0%, 80%) | $28^{th}$ May – $2^{nd}$ Jun | $28^{th}$ May – $2^{nd}$ Jun | $28^{th}$ May – $2^{nd}$ Jun | $28^{th}$ May – $2^{nd}$ Jun |
| | (2%,90%) | $9^{th}$ – $15^{th}$ April | $9^{th}$ – $15^{th}$ April | $7^{th}$ – $13^{th}$ May | $14^{th}$ – $20^{th}$ May |
| | (4%,80%) | $9^{th}$ – $15^{th}$ April | $9^{th}$ – $15^{th}$ April | $9^{th}$ – $15^{th}$ April | $7^{th}$ – $13^{th}$ May |
| | (10%,90%) | $9^{th}$ – $15^{th}$ April | $9^{th}$ – $15^{th}$ April | $9^{th}$ – $15^{th}$ April | $9^{th}$ – $15^{th}$ April |
| | (10%, 50%) | $9^{th}$ – $15^{th}$ April | $9^{th}$ – $15^{th}$ April | $9^{th}$ – $15^{th}$ April | $9^{th}$ – $15^{th}$ April |
| **Change in attack rates (times)** | (0, 100%) | 3.03 (0.26,11.29) | 3.94 (0.24,14.43) | 1.93 (0.07,5.64) | 2.16 (0.16,3.68) |
| | (0%, 90%) | 2.45 (0.27, 8.94) | 3.61 (0.21, 13.20) | 1.84 (0.07, 5.03) | 2.05 (0.16, 3.36) |
| | (2%,90%) | 1.87 (0.23, 6.60) | 1.72 (0.18, 6.46) | 0.71 (0.06,2.31) | 0.68 (0.09, 1.44) |
| | (4%,80%) | 1.26 (0.20, 4.09) | 1.06 (0.17, 3.48) | 0.41 (0.05,1.22) | 0.36 (0.07, 0.75) |
| | (10%,90%) | 0.96 (0.16, 2.62) | 0.77 (0.13, 2.21) | 0.27 (0.04,0.73) | 0.22 (0.05 0.45) |
| | (10%,50%) | 0.56 (0.15, 1.43) | 0.45 (0.12,1.20) | 0.16 (0.04,0.41) | 0.13 (0.04, 0.26) |

#### 8. Supplementary figures

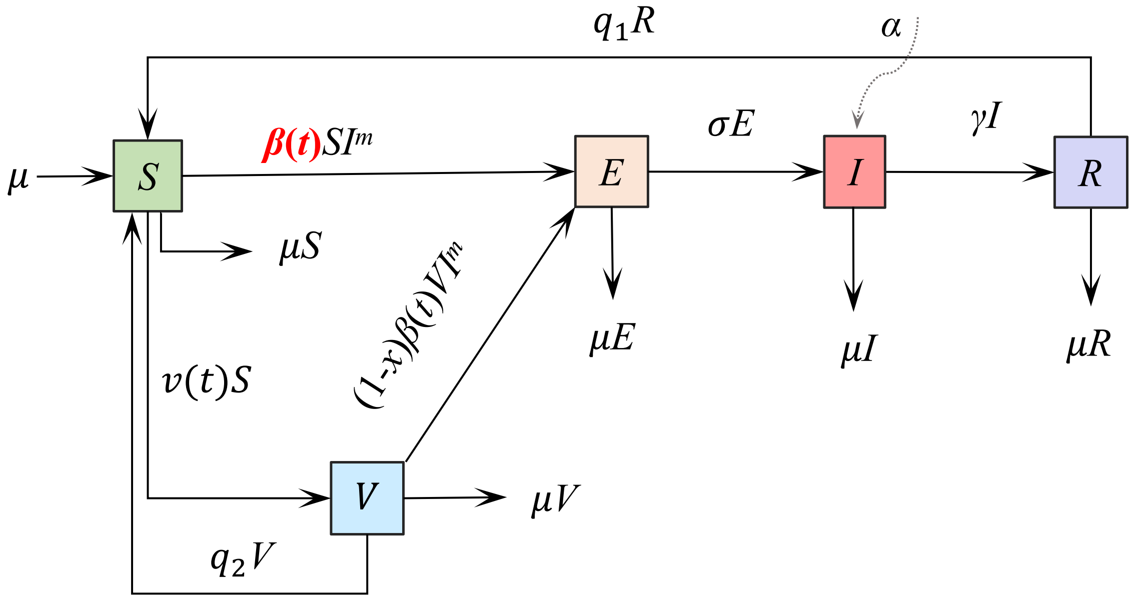

**Figure S1. The state-space mechanistic SVEIRS for influenza transmission where S, V, E, I, and R represented the compartments for the susceptible, vaccinated, exposed, infectious, and recovered individuals, respectively.** The descriptions of model parameters were shown in Table S1.

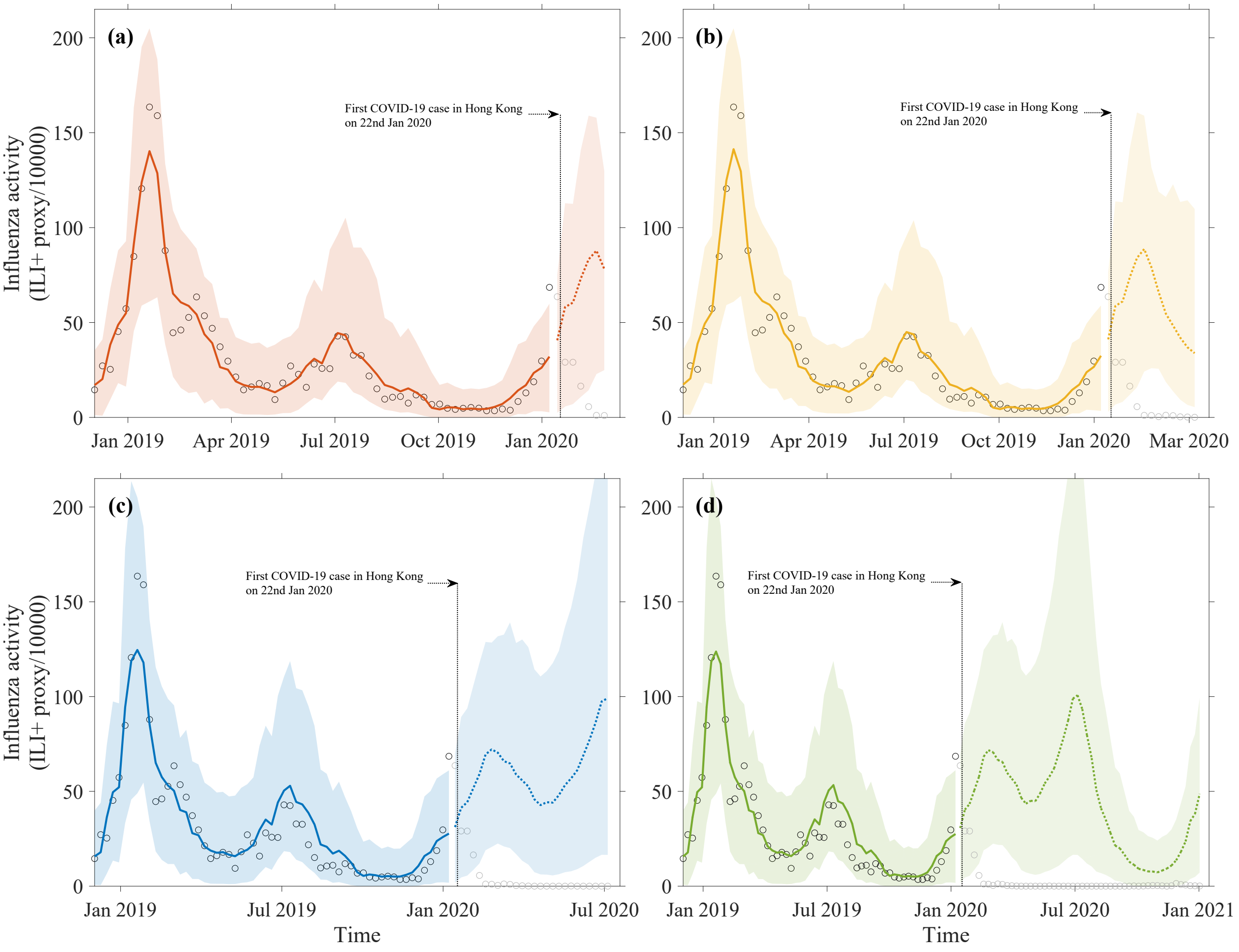

**Figure S2. The forecast of influenza activity in Hong Kong starting from 15th January 2020, for different forecasting windows: (a) 7 weeks ahead, (b) 13 weeks ahead, (c) 26 weeks ahead, and (d) 52 weeks ahead.** The black dots represent the observed influenza activities (ILI+ proxy/104) and the solid lines in respective colors represent the model predictions with 95% prediction intervals in shades. The dashed lines represent the mean of ensemble forecasts, and the colored shades represent their corresponding 95% prediction intervals.

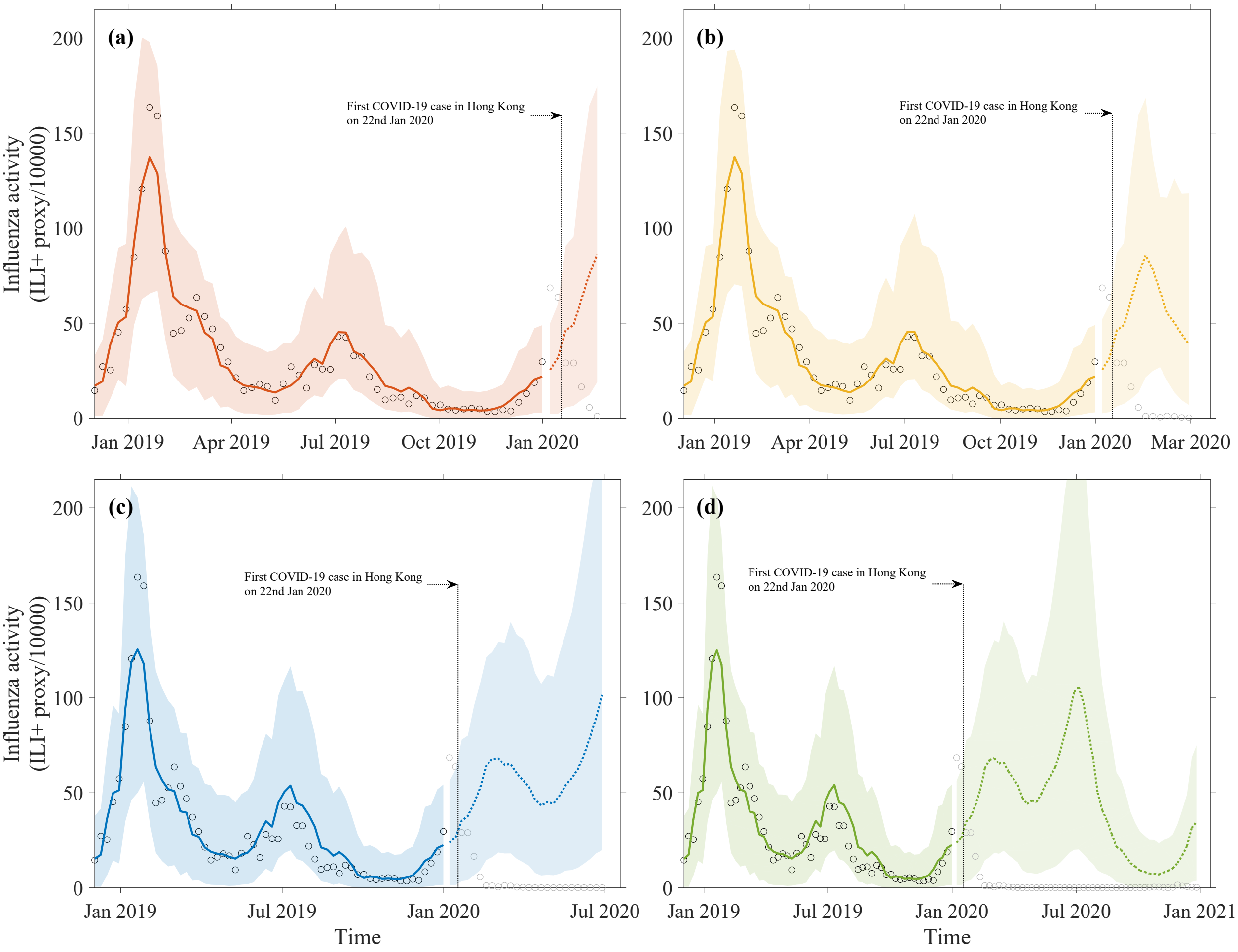

**Figure S3. The forecast of influenza activity in Hong Kong starting from 11th January 2020, for different forecasting windows: (a) 7 weeks ahead, (b)13 weeks ahead, (c) 26 weeks ahead, and (d) 52 weeks ahead.** The black circles represent the observed influenza activities (ILI+ proxy/$10^4$) and the solid lines in respective colors represent the model predictions with 95% prediction intervals in shades. The dashed lines represent the mean of ensemble forecasts, and the colored shades represent their corresponding 95% prediction intervals.

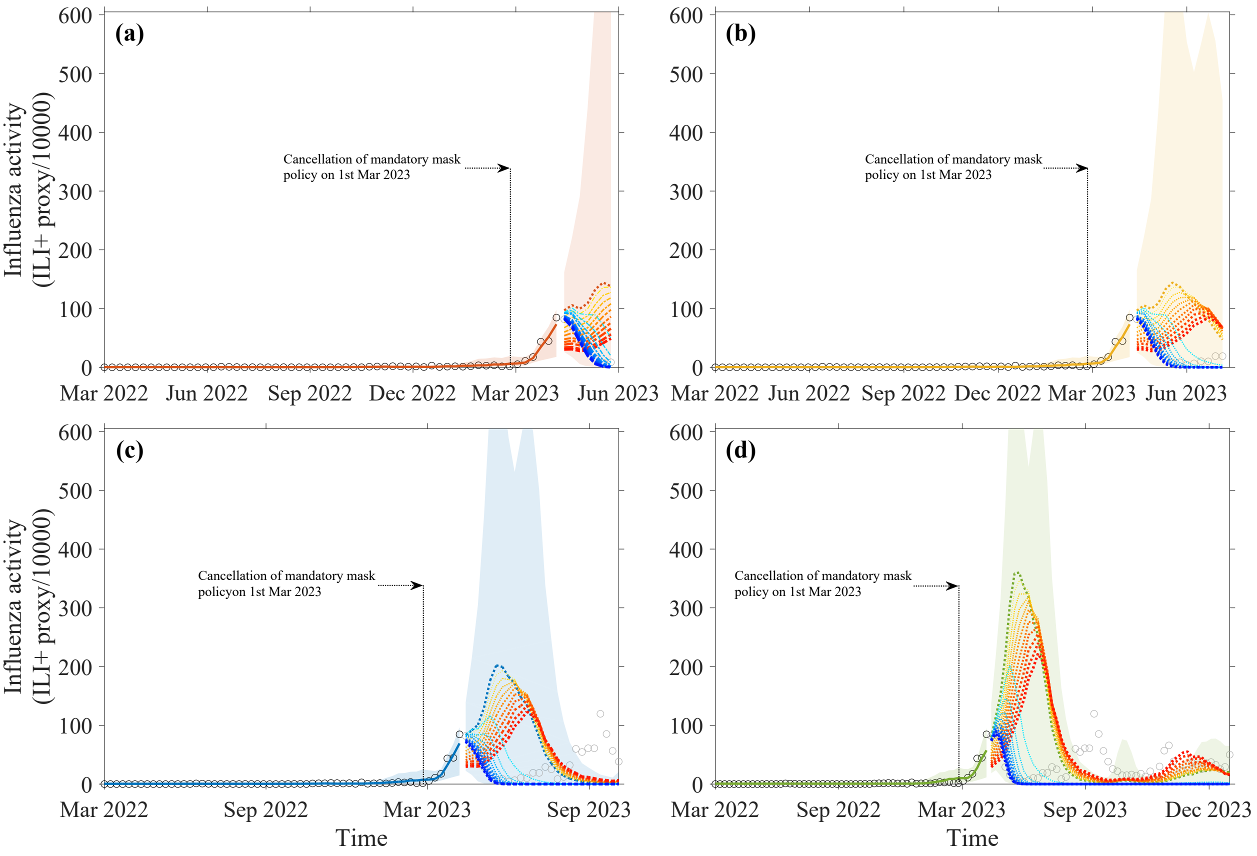

**Figure S4. The counterfactual forecasting and projection for 2023/24 season under varying levels of PHSMs and COVID-19-induced cross- protection(immunity/competition).** The baseline scenario (no cross-protection-induced reduction in susceptibility and complete removal of PHSMs, i.e., (0%, 100%)) is displayed in the same color as the model fit. The red lines represent different levels of residual impact of PHSMs in community (0%: orange, 100%: red); and blue lines represent diverse levels of COVID-19 driven cross-protection (0%: green, 10% darkblue).

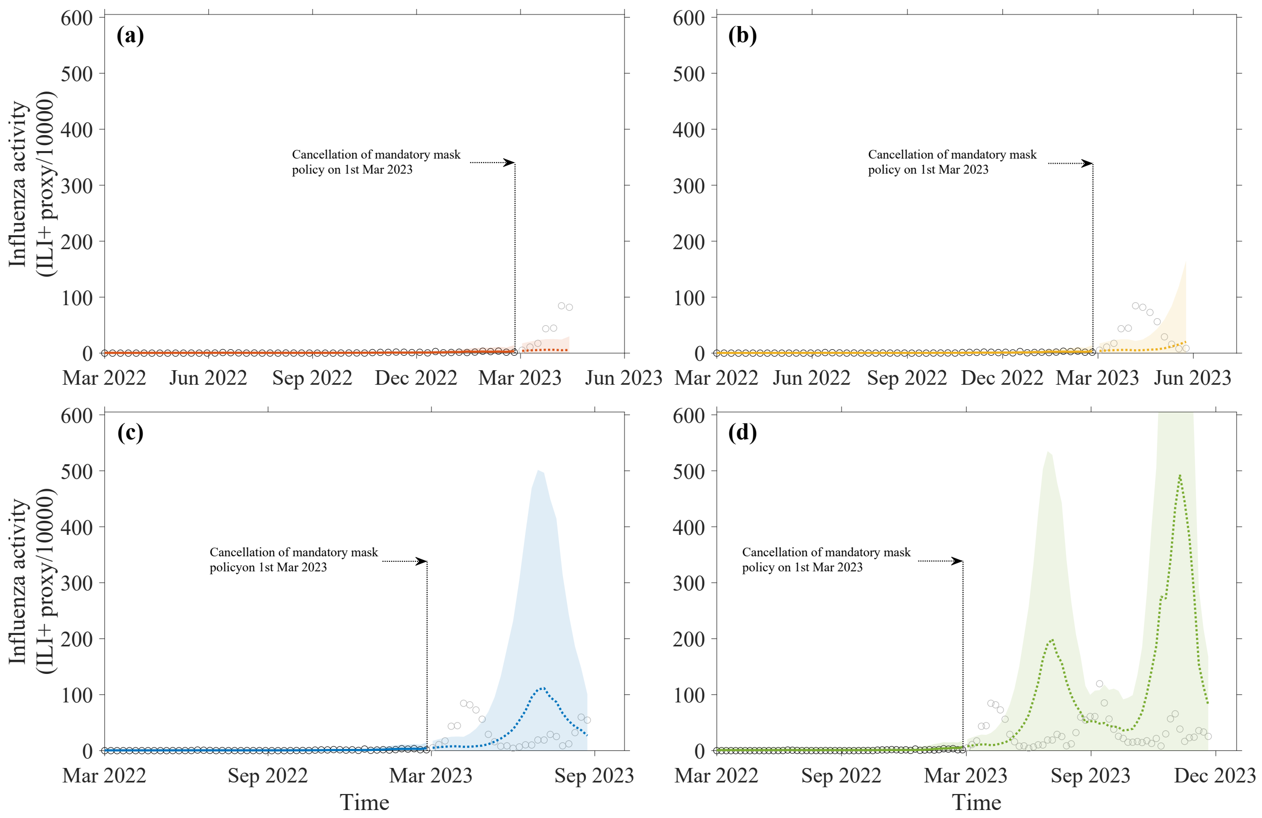

**Figure S5. The forecast of influenza activity starting from the 1st March 2023, for different periods: (a) 7 weeks ahead, (b) 13 weeks ahead, (c) 26 weeks ahead, and (d) 52 weeks ahead.** The black line represents the observed influenza activities (ILI+ proxy/$10^4$), and the dashed line represents the forecasted influenza activity, with colored shades indicating the 95% prediction intervals.

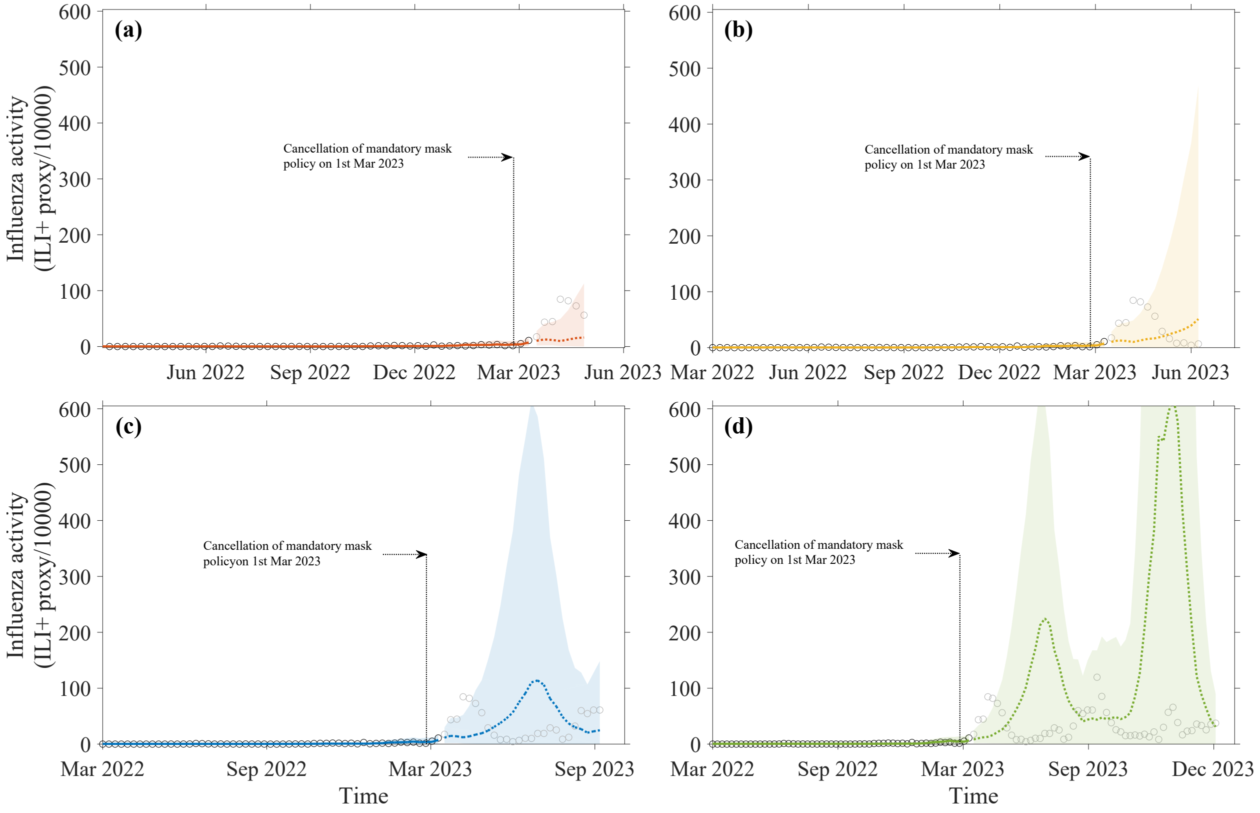

**Figure S6. The forecast of influenza activity starting from the 15th March 2023, for different periods: (a) 7 weeks ahead, (b) 13 weeks ahead, (c) 26 weeks ahead, and (d) 52 weeks ahead.** The black line represents the observed influenza activities (ILI+ proxy/$10^4$), and the dashed line represents the forecasted influenza activity, with colored shades indicating the 95% prediction intervals.

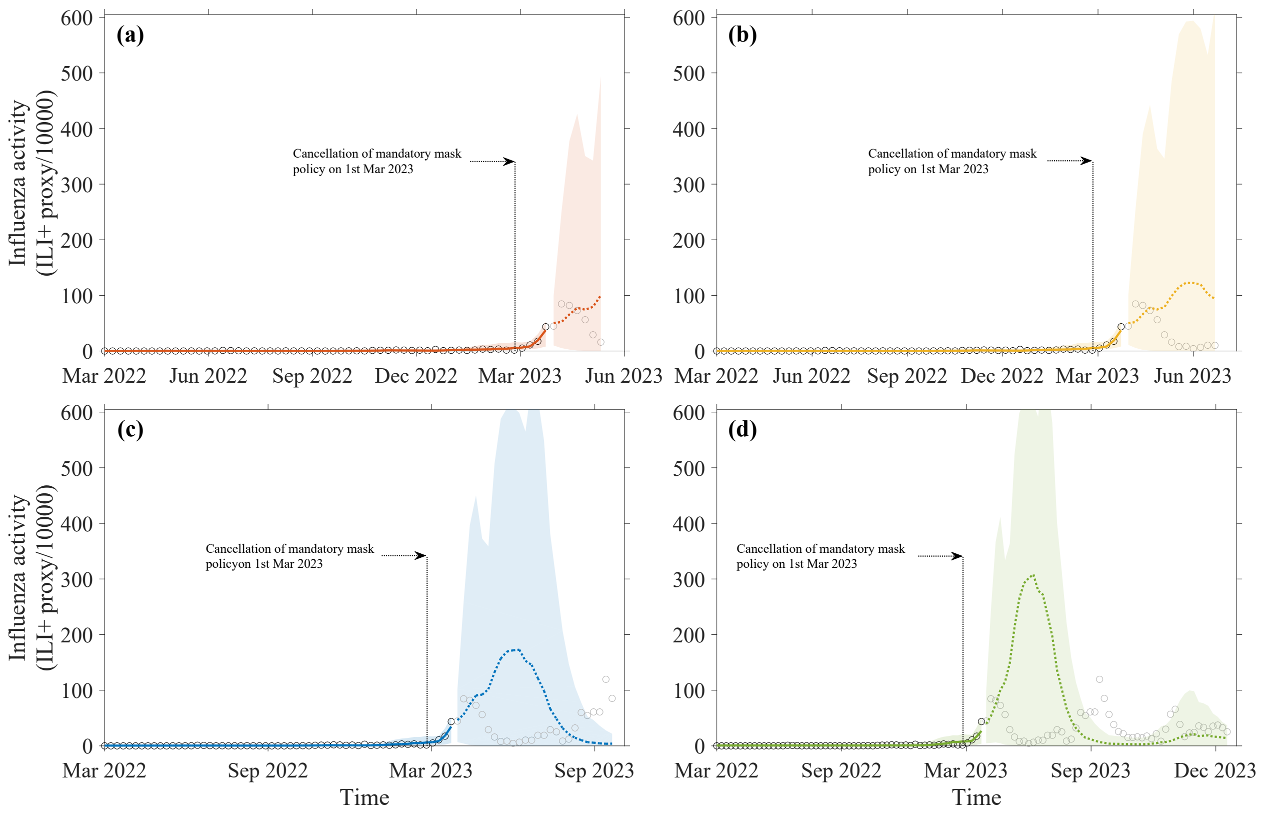

**Figure S7. The forecast of influenza activity starting from the 19th March2023, for different periods: (a) 7 weeks ahead, (b) 13 weeks ahead, (c) 26 weeks ahead, and (d) 52 weeks ahead.** The black line represents the observed influenza activities (ILI+ proxy/$10^4$), and the dashed line represents the forecasted influenza activity, with colored shades indicating the 95% prediction intervals.

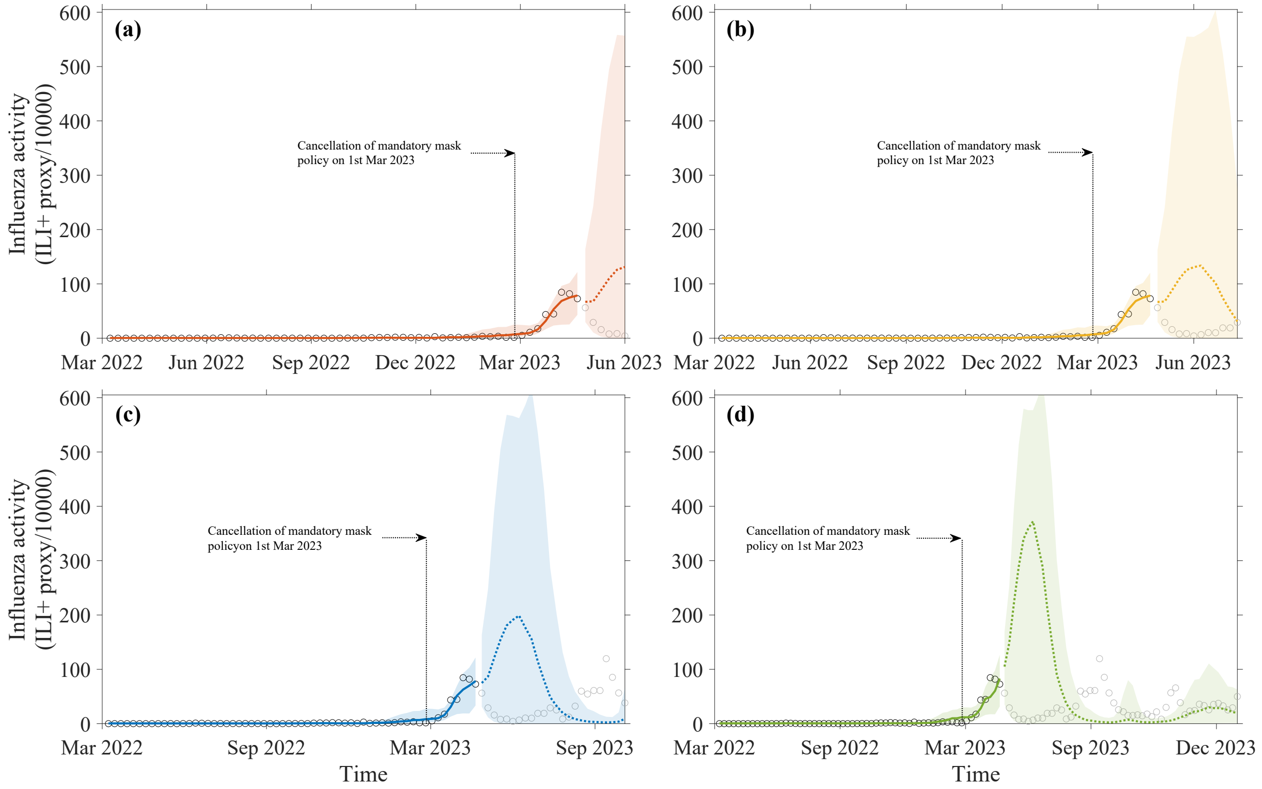

**Figure S8. The forecast of influenza activity starting from the 26th April 2023, for different periods: (a) 7 weeks ahead, (b) 13 weeks ahead, (c) 26 weeks ahead, and (d) 52 weeks ahead.** The black line represents the observed influenza activities (ILI+ proxy/$10^4$), and the dashed line represents the forecasted influenza activity, with colored shades indicating the 95% prediction intervals.

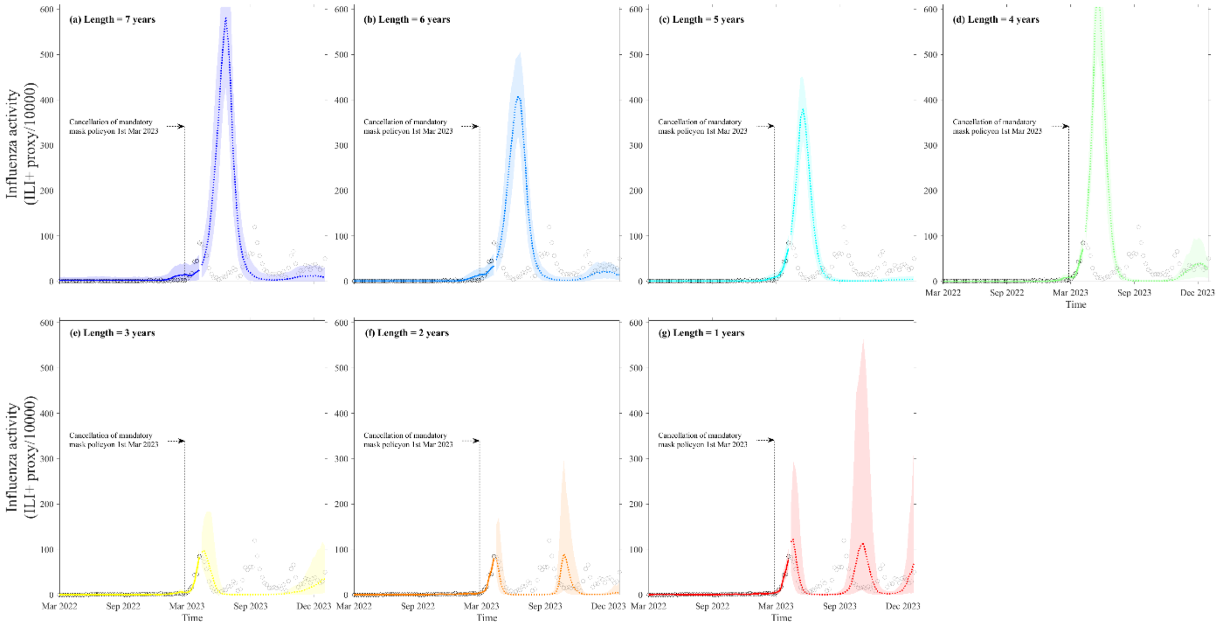

**Figure S9. The counterfactual forecasting for 2023/24 season in the post-pandemic era based on the optimal ensemble model by considering different lengths of historical data, (a)-(g) from 7 years to 1 years.** The black circle denoted the observed influenza activity (ILI+ proxy/$10^4$). The colored solid and dotted lines denote the fitting and forecasting, respectively. The colored shades denote their corresponding 95% prediction intervals.
